# Underlying cardiovascular risk and major adverse cardiovascular events after acute respiratory infection: a population-based cohort study of over 4.2 million individuals in England, 2008-2018

**DOI:** 10.1101/2021.03.18.21253890

**Authors:** Jennifer A Davidson, Amitava Banerjee, Liam Smeeth, Helen I McDonald, Daniel Grint, Emily Herrett, Harriet Forbes, Richard Pebody, Charlotte Warren-Gash

**Author notes:** **Correspondence to**: Jennifer Davidson, Department of Non-Communicable Disease Epidemiology, London School of Hygiene and Tropical Medicine, Keppel Street, London, WC1E 7HT, United Kingdom.

## Abstract

**Background:** While acute respiratory infections (ARIs) can lead to cardiovascular complications, the effect of underlying cardiovascular risk profile on ARI incidence and cardiovascular complications in those without established cardiovascular disease (CVD) is unknown. Whether to consider individuals at raised cardiovascular risk a priority group for vaccination against respiratory infections therefore remains unclear.

**Methods:** We conducted a cohort study in individuals aged 40-64 years without established CVD or a chronic health condition eligible for influenza vaccination, using Clinical Practice Research Datalink GOLD and Aurum data from 01/09/2008-31/08/2018 linked to Hospital Episode Statistics Admitted Patient Care and Office for National Statistics mortality data from England. We classified cardiovascular risk based on diagnosed hypertension and overall predicted cardiovascular risk estimated using QRISK2 score (≥10% compared with <10%). Using multivariable Poisson regression models, we obtained incidence rate ratios (IRR) for ARI. Among individuals who had an ARI, we then used multivariable Cox regression to obtain hazard ratios (HR) for the risk of major adverse cardiovascular events (MACE) within one year of infection.

**Findings:** 4,212,930 individuals were included; 12·5% had hypertension and 14·4% had a QRISK2 score ≥10%. After adjusting for confounders, patients with hypertension (IRR 1·04, 95% CI 1·03-1·05) or QRISK2 score ≥10% (IRR 1·39, 1·37-1·40) had a higher incidence of ARI. Of the 442,408 individuals with an ARI, 4,196 had a MACE within one year of infection. After adjustment, hypertension (HR 1·98, 1·83-2·15) and QRISK2 score ≥10% (HR 3·65, 3·42-3·89) were associated with substantial increased risk of a MACE after infection.

**Interpretation:** People without diagnosed CVD but who have raised cardiovascular risk, measured by diagnosed hypertension or, in particular, overall predicted cardiovascular risk, have increased incidence of both ARI and cardiovascular complications following an ARI.

## INTRODUCTION

The coronavirus (COVID-19) pandemic has expedited research on the role of acute respiratory infections (ARIs) in triggering cardiovascular complications. Before the pandemic, multiple observational studies identified ARIs increased the risk of myocardial infarction (MI) and stroke. In self-controlled case series (SCCS) studies, using large electronic health record (EHR) datasets, MI and stroke risks were elevated two- to six-fold in the days following clinically-diagnosed ARI and influenza-like illness (ILI), with risk remaining for up to one month.^1,2^ Laboratory-confirmed organisms, including *Streptococcus pneumonia* and influenza virus, cause consistent cardiovascular triggering effects.^3,4^

Observational studies and the few randomized controlled trials (RCTs) available show pneumococcal and influenza vaccines reduce the incidence of cardiovascular complications.^5–7^ A meta-analysis of secondary prevention RCTs found influenza vaccination reduced cardiovascular mortality by 55%.^7^ While not as widely researched, with no RCT evidence, a meta-analysis of observational studies in individuals ≥65 years found pneumococcal vaccination reduced the odds of acute coronary syndrome (ACS) by 17%.^5^

People without established cardiovascular disease (CVD) but with underlying cardiovascular risk are not typically recommended to receive seasonal influenza or pneumococcal vaccines by organizations such as the World Health Organization, the European Centre for Disease Prevention and Control, or in England, Public Health England. Public Health England recommends one-time polysaccharide pneumococcal vaccine and seasonal influenza vaccines for adults aged ≥65 years or <65 years with specific underlying health conditions, including those with chronic heart disease.^8^

Cohort studies have previously demonstrated an association between high blood pressure and cardiovascular complications in individuals as young as 40.^9,10^ Although blood pressure is an essential predictor of cardiovascular risk, it is only one component. Cardiovascular risk scores are increasingly used to predict the individual likelihood, most commonly 10-year risk, of future CVD based on multiple factors. QRISK2, now updated to a newer version called QRISK3, is embedded in UK primary care clinical management systems.^11^ The National Institute for Health and Care Excellence recommends QRISK2 for CVD risk assessment and management.^12^

Our study aimed to investigate how raised cardiovascular risk, defined by diagnosed hypertension and QRISK2 score, modified the occurrence of ARI and major adverse cardiovascular events (MACE) after an ARI using ten years of linked EHR data from England. A better understanding of risk factors for severe outcomes after ARI will inform stratified prevention and treatment strategies.

## METHODS

### Data sources

We conducted a retrospective cohort study using anonymized coded primary care data from the Clinical Practice Research Datalink (CPRD) databases, GOLD and Aurum.^13,14^ The databases comprise >35 million individuals, representative of the UK population in terms of age, sex and ethnicity. The coded data collected include: diagnoses, prescriptions, immunizations, and basic demographics.

Consenting practices have patient records linked to other data sources. We used linked secondary care data from the Hospital Episodes Statistics Admitted Patient Care (HES APC) database, Office of National Statistics (ONS) death data, and individual-level Townsend score deprivation twentiles. HES APC contains diagnoses made and procedures done for all NHS admissions in England coded using International Classification of Diseases 10th version (ICD-10).^15^ ONS deaths data, also coded using ICD-10, contains death certificates’ date and cause of death.

The CPRD Independent Scientific Advisory Committee approved data use for the study (application 19_209). CPRD provided relevant HES and ONS variables and Townsend data for the study population. London School of Hygiene and Tropical Medicine provided ethical approval (application 17894). All code lists used in our analysis are available from DOI: *awaiting*. The corresponding author had full access to the study data.

### Study population and follow-up

Eligible individuals were 40-64 year-olds not already eligible for influenza vaccination, with current registration at a CPRD (GOLD or Aurum) contributing practice and linked HES APC data in England from 01/09/2008-31/08/2018. Our selected time period covered the duration of QRISK2 use. Starting follow-up in September corresponded to when primary care practices identify individuals eligible for the seasonal influenza vaccine. We selected 40-64 years to include those with a higher likelihood of raised cardiovascular risk and MACE but below the age of universal influenza vaccination. We defined start of follow-up as the latest of; 01/09/2008, 40th birthday or research standard CPRD date (12 months after current registration date in Aurum and latest of 12 months after the current registration date or the practice research standard date in GOLD).

We excluded individuals who at baseline had; established CVD, previous pneumococcal vaccine, influenza vaccine within the 12 months before baseline, or a chronic health condition eligible for influenza vaccination recorded in CPRD. We defined conditions eligible for influenza vaccination as; chronic liver disease, chronic respiratory disease, chronic kidney disease stage 3-5, diabetes, asplenia or other splenic dysfunction, chronic neurological conditions, morbid obesity, or an immunosuppressive condition^8^. Full definitions of exclusion criteria are presented in **supplementary material appendix 1**.

We used the whole study population to investigate the incidence of ARI. Inclusion eligibility was re-assessed during follow-up when new measures became available, with follow-up ending at the earliest of; CVD or a chronic condition development, pneumococcal or influenza vaccination, death, transfer out of practice, practice last data collection, 65th birthday, or 31/08/2018.

Within the study population, individuals with an ARI were then followed-up from diagnosis to investigate the incidence and risk of MACE after this infection. Follow-up for this study population ended at the earliest of; MACE diagnosis, death, transfer out of practice, practice last data collection, one year after ARI diagnosis or 31/08/2018.

Figure 1. outlines the two follow-up periods; for ARI outcomes among the whole study population and for MACE within one year of ARI.

**Figure 1.**
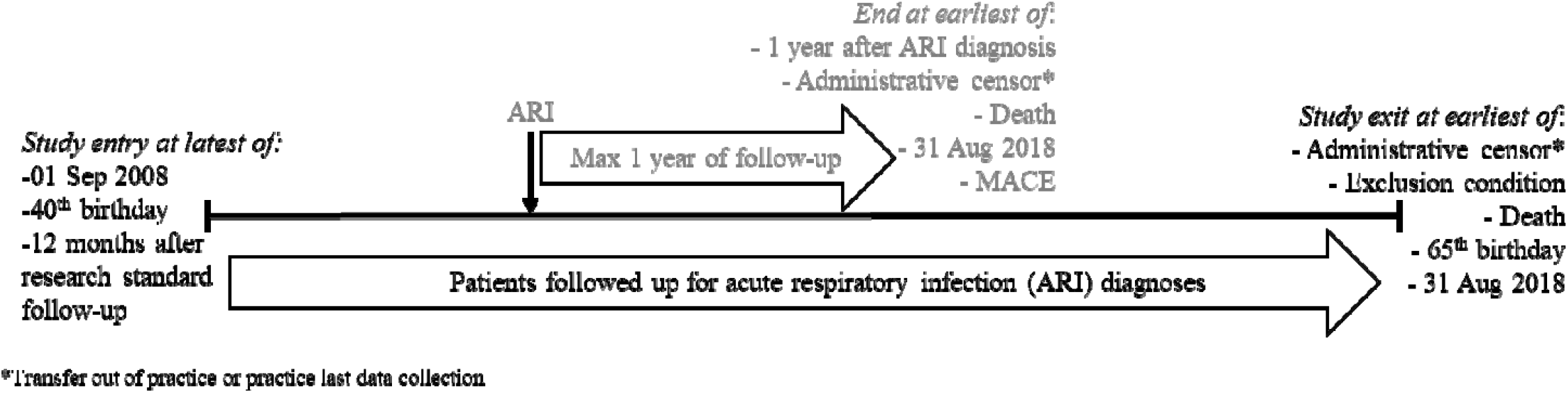
Study period and follow-up outline

### Acute respiratory infection identification

We defined ARI as a clinical or confirmed diagnosis of; pneumonia, acute bronchitis, influenza, influenza-like illness (ILI), or other acute infections suggestive of lower respiratory tract involvement.

We included diagnoses recorded in CPRD or HES APC, with selected codes based on previous codelists.^2^ In secondary analyses, we analyzed influenza/ILI and pneumonia separately. We grouped ARI records within 28 days of each other into a single episode. See **supplementary material appendix 2** for full details of episode definitions.

### Major adverse cardiovascular event identification

There is no single definition for the composite outcome of MACE. We used a broad definition of; ACS (MI and unstable angina), left ventricular heart failure, stroke or transient ischemic attack (TIA), acute limb ischemia, and cardiovascular death. We included diagnoses recorded in CPRD or HES APC, with the codes used informed by previous studies,^16^ and cardiovascular deaths (ICD-10 I00-I99) recorded in ONS data. Our secondary outcomes were each of the cardiovascular conditions separately.

### Cardiovascular risk identification

In separate analyses, we defined cardiovascular risk by hypertension or QRISK2 score. To ensure we only included individuals with persistent and diagnosed hypertension, we defined hypertension based on coded CPRD diagnoses with no time limit. The risk factors used in and process for the calculation of QRISK2 are outlined in **supplementary material appendix 3**.

Individuals were classified as having raised cardiovascular risk (hypertension or QRISK2 ≥10%) or not (no hypertension or QRISK2 <10%) at baseline. We updated cardiovascular risk when a new hypertension diagnosis was recorded or new measures relevant to the QRISK2 algorithm resulted in a score changing from <10% to ≥10%.

### Covariates

We considered demographics, lifestyle factors, and primary care consultation frequency in analyses of hypertension and ARI. The demographics were; age (5-year bands of 40-44, 45-49, 50-54, 55-59, and 60-64), sex (male and female), ethnicity (White, Black, South Asian, and Mixed/Other), and socioeconomic status (individual-level Townsend quintiles). The lifestyle factors were baseline; alcohol consumption, smoking status and BMI (underweight, normal, overweight or obese). Consultation frequency was derived based on the number of in-person or telephone consultations in the year before baseline. For QRISK2, we only included alcohol consumption and consultation frequency, as all other factors were in the QRISK2 algorithm. We identified included covariates using CPRD data, with HES data additionally used to identify ethnicity.

Our analyses of MACE outcomes after ARI again included the previously mentioned demographic and lifestyle factors. We stratified results by statin, antihypertensive or antiplatelet prescriptions in the year before infection.

### Statistical analysis

We pooled individual-level data from CPRD GOLD and Aurum.

By cardiovascular risk, we calculated crude annual age-stratified ARI incidence rates. We used Poisson regression to obtain incidence rate ratios (IRRs). Our random-effects models, to account for multiple episodes per individual, included time-updated exposure by age and cardiovascular risk. Initially, we adjusted for age and sex, then additional covariates. To assess the impact of ARI on the incidence and risk of MACE, we used Cox proportional hazards regression finely adjusted for calendar time. We used robust standard errors and initially adjusted for age and sex, then additional covariates. In all analyses, for individuals who entered and exited follow-up on the same day, we added one day to ensure all individuals contributed at least one day of follow-up.

We conducted all analyses in Stata (version 16).

### Sensitivity analyses

We conducted five pre-specified sensitivity analyses. (i) We validated the results obtained from combined individual-level GOLD and Aurum data by analyzing each separately and combining them with random-effect meta-analysis. We assessed between-database heterogeneity with the I2 statistic. (ii) We only excluded individuals eligible for both pneumococcal and influenza vaccination,^8^ rather than the broader influenza vaccine recommendations used in our main analyses (see **supplementary material appendix 1**). (iii) To validate our QRISK2 algorithm, we repeated analyses restricted to individuals with a QRISK2 score recorded in CPRD from 2015-2017 (see **supplementary material appendix 3**). (iv) We used a narrower MACE definition with only the most severe outcomes; MI, heart failure, stroke and cardiovascular death. (v) We repeated our MACE after ARI analysis without individuals who received pneumococcal or influenza vaccine during follow-up.

## RESULTS

### Description of the cohort

The study cohort included 4,212,930 individuals (773,362 from GOLD and 3,439,568 from Aurum) with a median follow-up of 3·9 (IQR=1·6-7·6) years (**figure 2**). Among the cohort, 52·9% were men and the median age at baseline was 46 (IQR=40-53) years (**table 1**). The GOLD and Aurum-recorded individuals’ demographic and lifestyle characteristics were similar, except higher proportions of Aurum-recorded individuals were non-White, resident in the most deprived geographies and current or ex-smokers (supplementary materials table S1).

**Figure 2.**
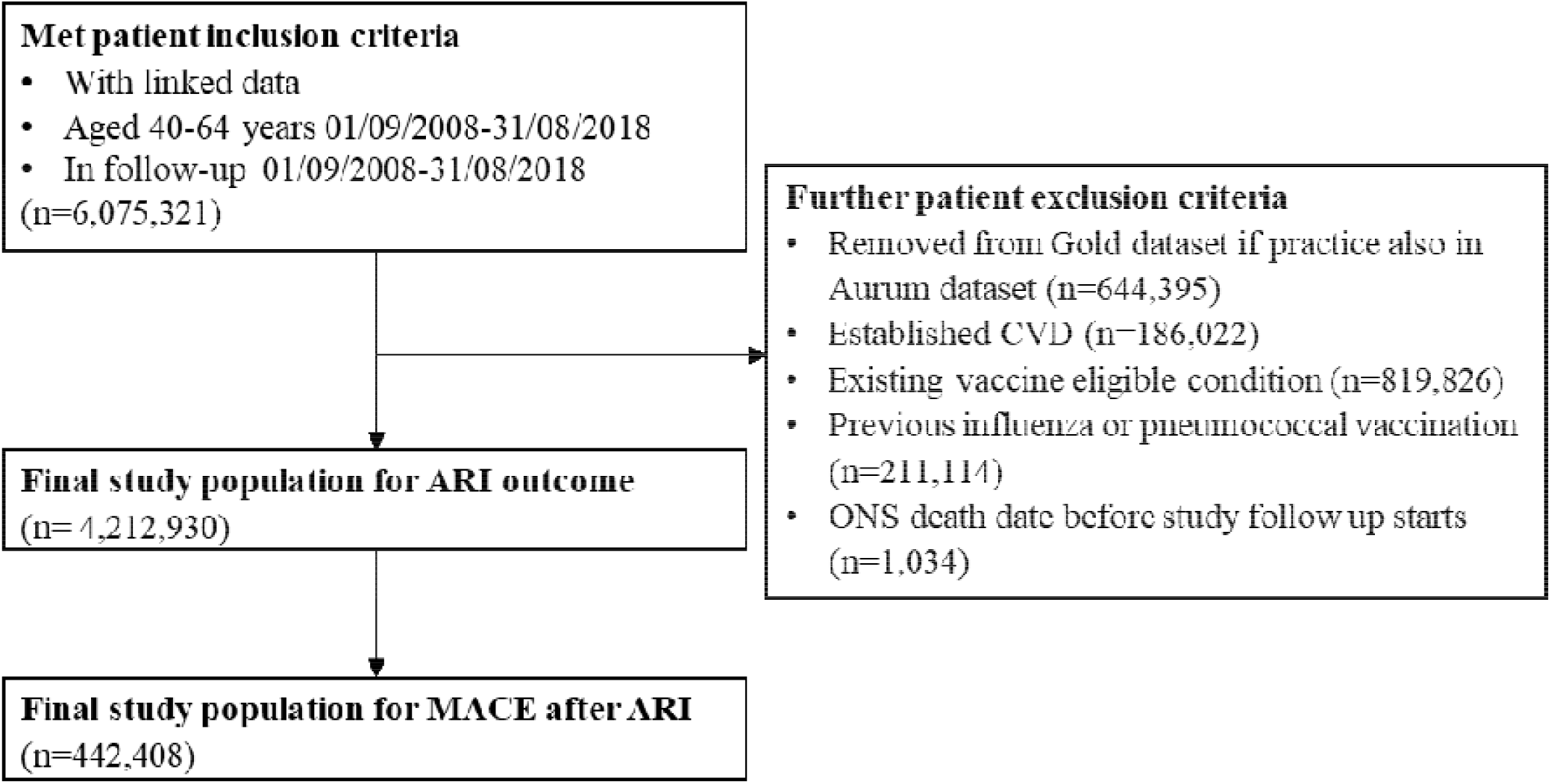
Study population inclusion flowchart

**Table 1.** Baseline demographic and lifestyle characteristics of included study population

|  | Cohort<br>n = 4,212,930 |
| --- | --- |
| Age (years) |  |
| 40-44 | 1,920,369 (45.6%) |
| 45-49 | 782,897 (18.6%) |
| 50-54 | 612,202 (14.5%) |
| 55-59 | 490,619 (11.6%) |
| 60-64 | 406,843 (9.7%) |
| Sex (n = 4,212,898) |  |
| Male | 2,226,561 (52.9%) |
| Female | 1,986,337 (47.1%) |
| Ethnicity (n = 3,702,718) |  |
| White | 3,242,107 (87.6%) |
| South Asian | 194,931 (5.3%) |
| Black | 154,270 (4.2%) |
| Mixed/Other | 111,410 (3.0%) |
| Townsend quintile (n = 4,207,605) |  |
| 1 (least deprived) | 1,004,670 (23.9%) |
| 2 | 905,691 (21.5%) |
| 3 | 825,679 (19.6%) |
| 4 | 739,187 (17.6%) |
| 5 (most deprived) | 732,378 (17.4%) |
| BMI category* (n = 3,447,604) |  |
| Underweight (<18.5 kg/m<sup>2</sup>) | 51,002 (1.5%) |
| Normal weight (18.5-24.9 kg/m<sup>2</sup>) | 1,449,683 (42.0%) |
| Overweight (25.0-29.9 kg/m<sup>2</sup>) | 1,274,701 (37.0%) |
| Obese (30.0-39.9 kg/m<sup>2</sup>) | 672,218 (19.5%) |
| Smoking status* (n = 4,082,791) |  |
| Non-smoker | 1,686,919 (41.3%) |
| Current smoker | 1,076,707 (26.4%) |
| Ex-smoker | 1,319,165 (32.3%) |
| Alcohol consumption* (n = 3,674,494) |  |
| No known heavy drinking | 3,477,336 (94.6%) |
| Heavy drinking | 197,158 (5.4%) |

Data are n (%). \*Closest measure before start of follow-up.

There were 526,480 (12·5%) individuals with diagnosed hypertension; 347,418 had a diagnosis at baseline and a further 179,062 during follow-up. Of the 607,087 (14·4%) individuals with QRISK2 ≥10%, 402,594 had this score at baseline and 204,493 had a score increase from <10% to ≥10% during follow-up. Overall, 239,184 individuals had both hypertension and QRISK ≥10%.

### Acute respiratory infections

There were 586,147 ARI episodes among 442,408 individuals; 107,639 episodes were influenza/ILI and 31,068 pneumonia. The rate of ARI and pneumonia increased with age, other than among individuals with ARI and QRISK2 ≥10% for whom there was no age trend (**figure 3**). The rate of influenza/ILI decreased with age. The incidence of ARI was higher among individuals with hypertension (40·3/1,000 person-years) and QRISK2 ≥10% (43·8/1,000 person-years), compared to those without hypertension (29·1/1,000 person-years) and QRISK2 <10% (28·8/1,000 person-years).

**Figure 3.**
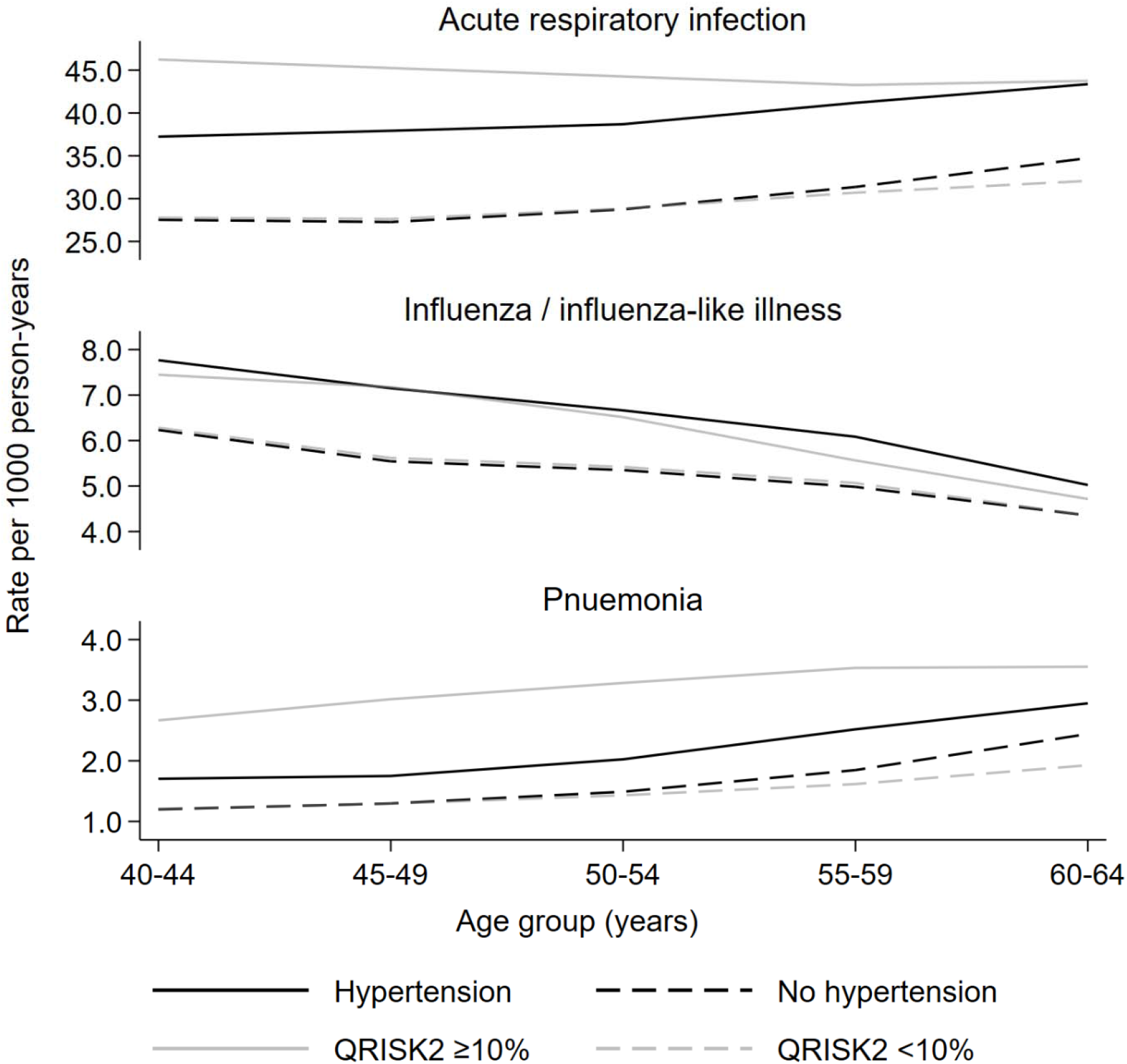
Age-specific infection rates by cardiovascular risk group

After adjustment for confounders, the association between hypertension and ARI was largely removed (IRR 1·04, 95% CI 1·03-1·05) but remained for QRISK2 ≥10% and ARI (IRR 1·39, 1·37-1·40) (**table 2**). The increased incidence of pneumonia among individuals with hypertension (IRR 1·12, 1·07-1·16) and QRISK ≥10% (IRR 2·32, 2·25-2·40) was more pronounced. Results differed for influenza/ILI, with a lower incidence among those with hypertension (IRR 0·98, 0·96-1·00) and QRISK2 ≥10% (IRR 0·88, 0·86-0·90).

**Table 2.** Crude and adjusted incidence rate ratios for the association between cardiovascular risk and acute respiratory infections

| Outcome | Cardiovascular risk | No. of events | Rate per 1,000 person-years | Crude IRR (95% CI) | Age and sex-adjusted IRR (95% CI) | Fully-adjusted* IRR (95% CI) |
| --- | --- | --- | --- | --- | --- | --- |
| Acute respiratory infection | Hypertension | 77,674 | 40.3 (40.0-40.7) | 1.38 (1.36-1.39) | 1.33 (1.32-1.34) | 1.04 (1.03-1.05) |
|  | No hypertension | 508,473 | 29.1 (29.0-29.2) | 1 | 1 | 1 |
| | QRISK2 $\geq 10\%$ | 81,662 | 43.8 (43.4-44.2) | 1.52 (1.50-1.53) | - | 1.39 (1.37-1.40) |
| | QRISK2 $< 10\%$ | 504,485 | 28.8 (28.7-28.9) | 1 | - | 1 |
| Influenza/ILI | Hypertension | 12,050 | 6.3 (6.1-6.4) | 1.14 (1.11-1.16) | 1.25 (1.22-1.27) | 0.98 (0.96-1.00) |
|  | No hypertension | 95,589 | 5.5 (5.4-5.5) | 1 | 1 | 1 |
| | QRISK2 $\geq 10\%$ | 10,010 | 5.4 (5.3-5.5) | 0.96 (0.94-0.98) | - | 0.88 (0.86-0.90) |
| | QRISK2 $< 10\%$ | 97,629 | 5.6 (5.5-5.6) | 1 | - | 1 |
| Pneumonia | Hypertension | 4,479 | 2.3 (2.2-2.4) | 1.59 (1.53-1.65) | 1.32 (1.27-1.38) | 1.12 (1.07-1.16) |
|  | No hypertension | 26,589 | 1.5 (1.5-1.5) | 1 | 1 | 1 |
| | QRISK2 $\geq 10\%$ | 6,476 | 3.5 (3.4-3.6) | 2.60 (2.52-2.69) | - | 2.32 (2.25-2.40) |
| | QRISK2 $< 10\%$ | 24,592 | 1.4 (1.4-1.4) | 1 | - | 1 |

Total person-years per 1,000: hypertension = 1,926.2 no hypertension = 17,467.9, QRISK2 $\geq 10\%$ = 1,865.1 and QRISK2 $< 10\%$ = 17,529.1 LRT p-values all $< 0.001$ . \*Hypertension models adjusted for: age, sex, ethnicity, socioeconomic status, BMI, alcohol intake, smoking status and consultation frequency. QRISK2 models adjusted for: alcohol intake and consultation frequency.

#### Sensitivity analyses

When we carried out the following sensitivity analyses, results were similar: (i) analyzed GOLD and Aurum separately, and combined results in meta-analysis (**supplementary material table S2**), (ii) redefined study population to exclude individuals with a chronic condition eligible for both influenza and pneumococcal vaccines (**supplementary material table S3**), and (iii) compared recorded and calculated QRISK2 (**supplementary material appendix 3**).

### Major adverse cardiovascular events after an acute respiratory infection

Among the 442,408 individuals (with 526,800 episodes) who had an ARI, 4,169 had a MACE within one year. Over one-third (38·5%) of the ARIs followed by a MACE were pneumonia.

There were 985 (11·2/1,000 person-years) MACE in individuals with hypertension, 3,184 (5·8/1,000 person-years) in those without hypertension, 1,526 (17·5/1,000 person-years) in individuals with QRISK2 ≥10% and 2,643 in those with QRISK2 <10% (4·8/1,000 person-years) (**table 3**). **Supplementary material tables S4 and S5** show results for MACE after influenza/ILI and pneumonia.

**Table 3.**
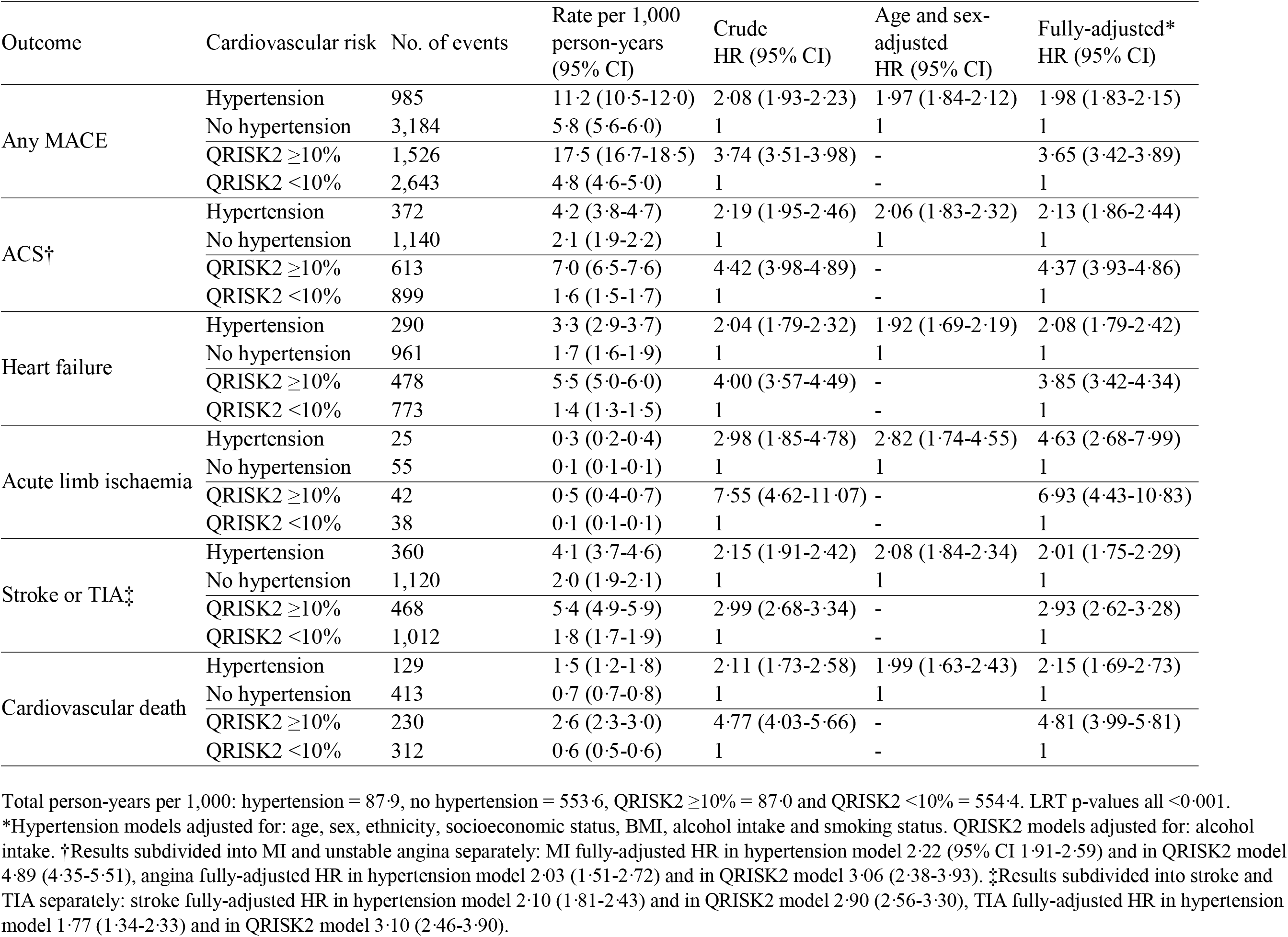
MACE after ARI incidence rates and incidence rate ratios by cardiovascular risk group

| Outcome | Cardiovascular risk | No. of events | Rate per 1,000 person-years (95% CI) | Crude HR (95% CI) | Age and sex-adjusted HR (95% CI) | Fully-adjusted* HR (95% CI) |
| --- | --- | --- | --- | --- | --- | --- |
| Any MACE | Hypertension | 985 | 11.2 (10.5-12.0) | 2.08 (1.93-2.23) | 1.97 (1.84-2.12) | 1.98 (1.83-2.15) |
|  | No hypertension | 3,184 | 5.8 (5.6-6.0) | 1 | 1 | 1 |
| | QRISK2 $\geq 10\%$ | 1,526 | 17.5 (16.7-18.5) | 3.74 (3.51-3.98) | - | 3.65 (3.42-3.89) |
| | QRISK2 $< 10\%$ | 2,643 | 4.8 (4.6-5.0) | 1 | - | 1 |
| ACS† | Hypertension | 372 | 4.2 (3.8-4.7) | 2.19 (1.95-2.46) | 2.06 (1.83-2.32) | 2.13 (1.86-2.44) |
|  | No hypertension | 1,140 | 2.1 (1.9-2.2) | 1 | 1 | 1 |
| | QRISK2 $\geq 10\%$ | 613 | 7.0 (6.5-7.6) | 4.42 (3.98-4.89) | - | 4.37 (3.93-4.86) |
| | QRISK2 $< 10\%$ | 899 | 1.6 (1.5-1.7) | 1 | - | 1 |
| Heart failure | Hypertension | 290 | 3.3 (2.9-3.7) | 2.04 (1.79-2.32) | 1.92 (1.69-2.19) | 2.08 (1.79-2.42) |
|  | No hypertension | 961 | 1.7 (1.6-1.9) | 1 | 1 | 1 |
| | QRISK2 $\geq 10\%$ | 478 | 5.5 (5.0-6.0) | 4.00 (3.57-4.49) | - | 3.85 (3.42-4.34) |
| | QRISK2 $< 10\%$ | 773 | 1.4 (1.3-1.5) | 1 | - | 1 |
| Acute limb ischaemia | Hypertension | 25 | 0.3 (0.2-0.4) | 2.98 (1.85-4.78) | 2.82 (1.74-4.55) | 4.63 (2.68-7.99) |
|  | No hypertension | 55 | 0.1 (0.1-0.1) | 1 | 1 | 1 |
| | QRISK2 $\geq 10\%$ | 42 | 0.5 (0.4-0.7) | 7.55 (4.62-11.07) | - | 6.93 (4.43-10.83) |
| | QRISK2 $< 10\%$ | 38 | 0.1 (0.1-0.1) | 1 | - | 1 |
| Stroke or TIA‡ | Hypertension | 360 | 4.1 (3.7-4.6) | 2.15 (1.91-2.42) | 2.08 (1.84-2.34) | 2.01 (1.75-2.29) |
|  | No hypertension | 1,120 | 2.0 (1.9-2.1) | 1 | 1 | 1 |
| | QRISK2 $\geq 10\%$ | 468 | 5.4 (4.9-5.9) | 2.99 (2.68-3.34) | - | 2.93 (2.62-3.28) |
| | QRISK2 $< 10\%$ | 1,012 | 1.8 (1.7-1.9) | 1 | - | 1 |
| Cardiovascular death | Hypertension | 129 | 1.5 (1.2-1.8) | 2.11 (1.73-2.58) | 1.99 (1.63-2.43) | 2.15 (1.69-2.73) |
|  | No hypertension | 413 | 0.7 (0.7-0.8) | 1 | 1 | 1 |
| | QRISK2 $\geq 10\%$ | 230 | 2.6 (2.3-3.0) | 4.77 (4.03-5.66) | - | 4.81 (3.99-5.81) |
| | QRISK2 $< 10\%$ | 312 | 0.6 (0.5-0.6) | 1 | - | 1 |

Total person-years per 1,000: hypertension = 87.9, no hypertension = 553.6, QRISK2 $\geq 10\%$ = 87.0 and QRISK2 $< 10\%$ = 554.4. LRT p-values all $< 0.001$ .
\*Hypertension models adjusted for: age, sex, ethnicity, socioeconomic status, BMI, alcohol intake and smoking status. QRISK2 models adjusted for: alcohol intake. †Results subdivided into MI and unstable angina separately: MI fully-adjusted HR in hypertension model 2.22 (95% CI 1.91-2.59) and in QRISK2 model 4.89 (4.35-5.51), angina fully-adjusted HR in hypertension model 2.03 (1.51-2.72) and in QRISK2 model 3.06 (2.38-3.93). ‡Results subdivided into stroke and TIA separately: stroke fully-adjusted HR in hypertension model 2.10 (1.81-2.43) and in QRISK2 model 2.90 (2.56-3.30), TIA fully-adjusted HR in hypertension model 1.77 (1.34-2.33) and in QRISK2 model 3.10 (2.46-3.90).

Half (50·5%) of MACE occurred within 30 days of ARI (**figure 4**), but the median interval was slightly longer in individuals with hypertension (median=41 [IQR=1-168] days) or QRISK2 ≥10% (median=31 [IQR=1-60] days), compared to those without hypertension (median=24 [IQR=1-156] days) or QRISK2 <10% (median 26 [IQR=1-159] days). The immediacy of MACE after pneumonia was even more pronounced. In comparison, there was a greater spread in the timing of MACE after influenza/ILI.

**Figure 4.**
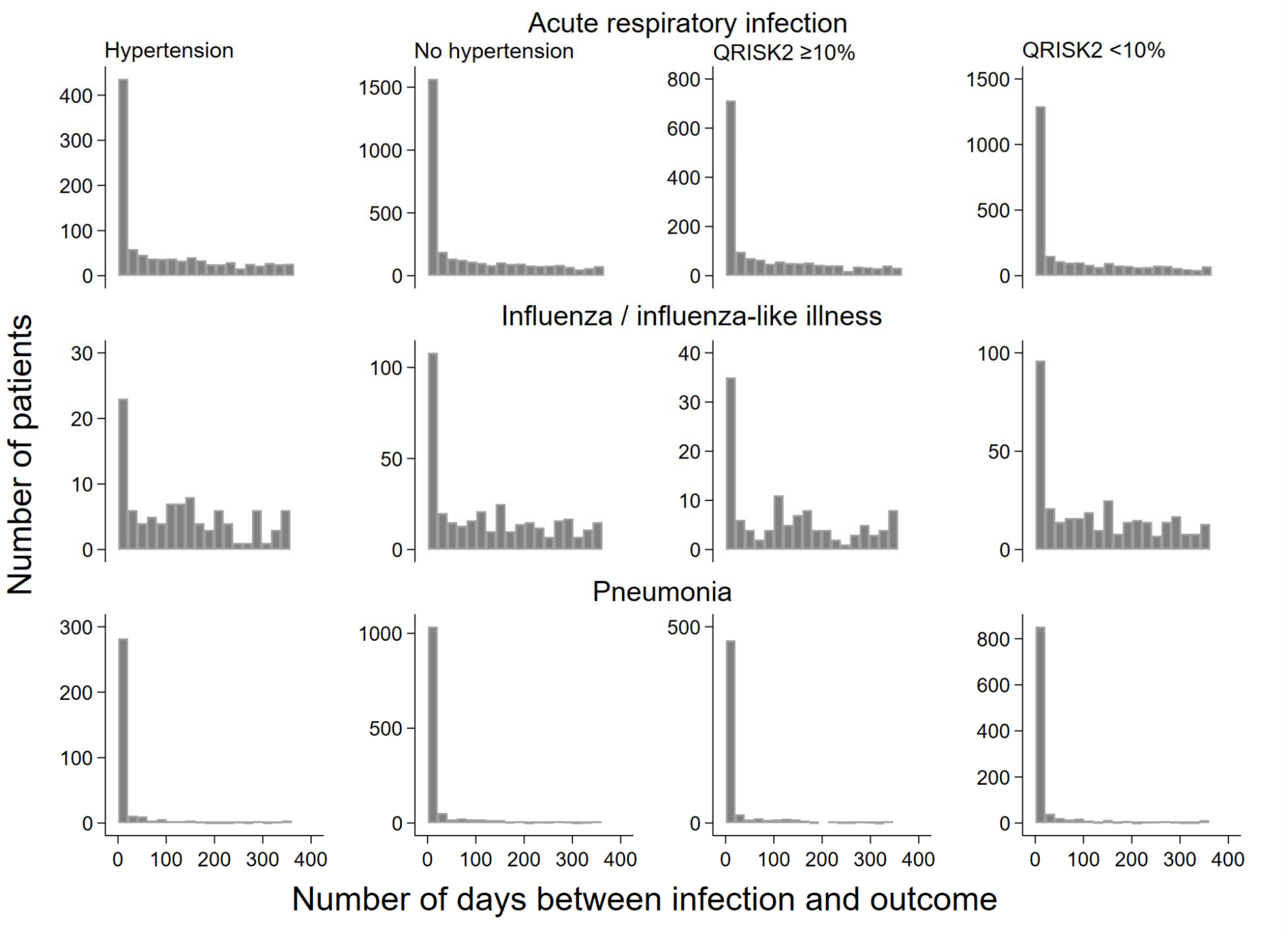
Timing between acute respiratory infection and major adverse cardiovascular event

After adjustment for confounders, the association between hypertension (hazard ratio [HR] 1·98, 1·83-2·15) or QRISK2 ≥10% (HR 3·65, 3·42-3·89) and MACE after ARI remained (**table 3**). For our secondary outcomes, the HRs were largest for acute limb ischaemia (hypertension: 4·63, 2·68-7·99 and QRISK2: 6·93, 4·43-10·83) – although there were only a small number of events, ACS (hypertension: 2·13, 1·86-2·44 and QRISK2: 4·37, 3·93-4·86), and cardiovascular death (hypertension: 2·15, 1·69-2·73 and QRISK2: 4·81, 3·99-5·81).

In analyses stratified by antihypertensive, statin and antiplatelet prescription, the association between raised cardiovascular risk and MACE after ARI was reduced among those with a prescription but remained increased by QRISK2 (**figure 5, supplementary material table S6**).

**Figure 5.**
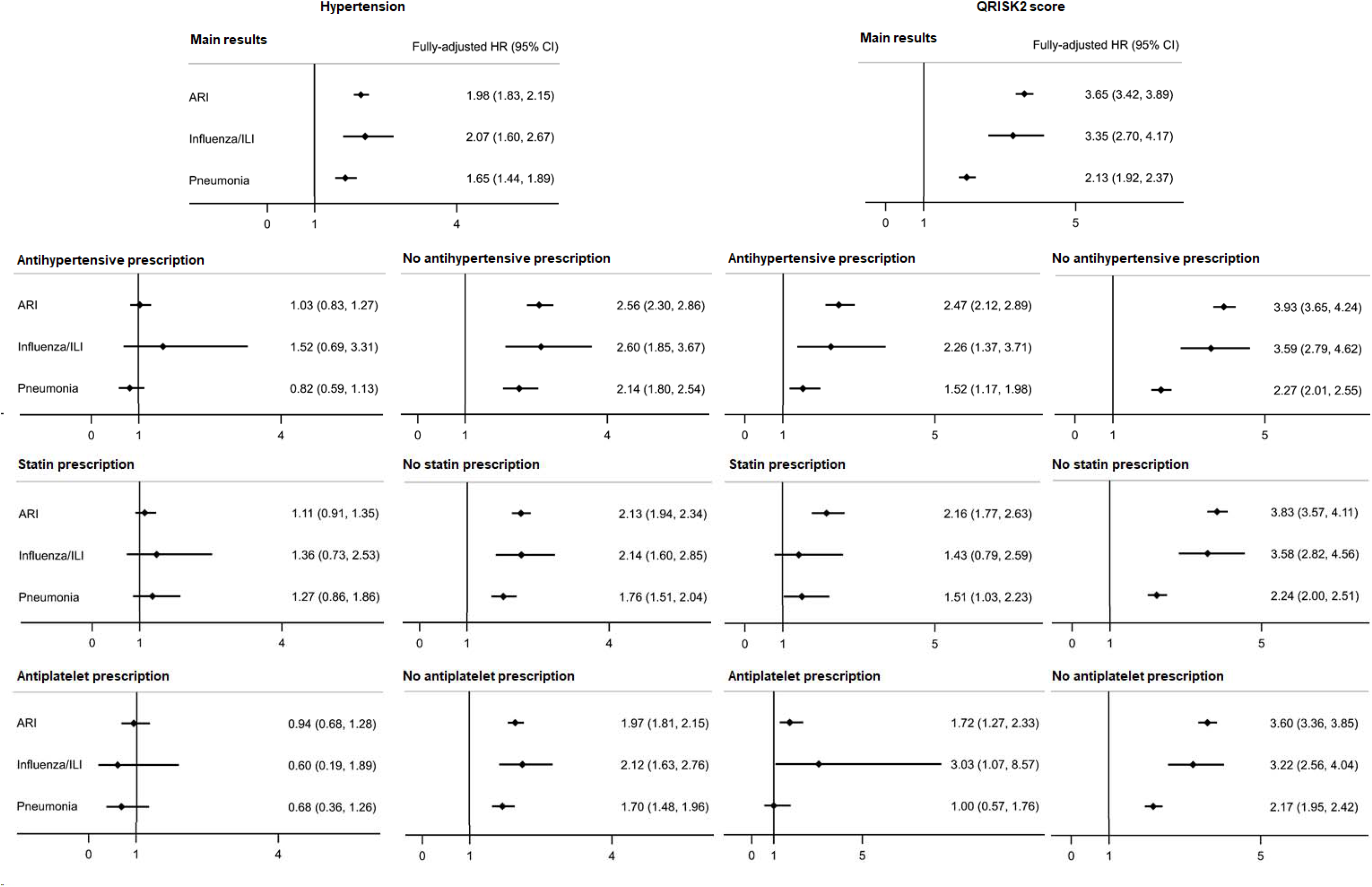
Fully-adjusted hazard ratios for major adverse cardiovascular event after acute respiratory infection by stratifying factor

#### Sensitivity analyses

When we carried out the following sensitivity analyses, results were similar overall: (i) analyzed CPRD GOLD and Aurum separately, and combined results in meta-analysis (**supplementary material table S7**), (ii) redefined study population to exclude individuals who have a chronic condition eligible for both influenza and pneumococcal vaccines (**supplementary material table S8**), (iii) compared recorded and calculated QRISK2 (**supplementary material appendix 3**), (iv) redefined outcome to only include severe MACE (**supplementary material table S9**), and (v) excluded individuals who received influenza or pneumococcal vaccine in follow-up (**supplementary material table S10**).

## DISCUSSION

### Summary

In our study population of >4·2 million adults 40-64 years with no chronic health condition eligible for influenza vaccination, individuals with raised cardiovascular risk measured by diagnosed hypertension or QRISK2 ≥10% had increased incidence of both ARI and MACE after ARI, compared with those at low cardiovascular risk. ARI incidence rates were 1·4- to 1·5-fold higher among individuals with raised cardiovascular risk. After adjustment for potential explanatory variables, the increase in incidence was marginal when risk was defined by hypertension diagnosis and more substantial when measured by QRISK2. The increase was more considerable among individuals who had pneumonia. We observed no difference in ILI incidence by hypertension and reduced ILI incidence among individuals with a QRISK2 ≥10%. Again, in the association between raised cardiovascular risk and MACE after ARI, we observed a substantially larger increase in risk by QRISK2 (3·7-fold) than hypertension (2-fold). Associations were similar for ARI overall, ILI and pneumonia. The median time between ARI and MACE was 24-41 days.

## Results in context

Our finding of increased ARI incidence among individuals with raised cardiovascular risk is consistent with a recent UK Biobank study which examined the relationship between blood pressure and risk of several different respiratory infections.^17^ Among the Biobank participants, prevalent hypertension was, after adjustment for possible explanatory factors, associated with increased risk of pneumonia (HR 1·36, 1·29-1·43), influenza or viral pneumonia (HR 1·12, 1·01-1·23), and other lower respiratory infections (HR 1·15, 1·11-1·19). In our study, the increase in ARI incidence among hypertensive individuals, measured by recorded diagnoses, was smaller. Instead, we found more substantial increases in those with QRISK2 ≥10%, which includes systolic blood pressure reading and antihypertensive use. Only 39% of patients with QRISK2 ≥10% had diagnosed hypertension. In other UK cohorts, influenza infection increases were associated with non-White ethnicity and deprivation.^18,19^ Both ethnicity and socioeconomic status are risk factors for CVD, and included in QRISK2.

Previous SCCS studies reported transient two- to six-fold increases in MI and stroke risk in the days after ARI.^1–4^ In the few observational studies to consider the impact of cardiovascular risk on this association, results were mixed. In two studies hypertension was associated with an increase in cardiovascular complications after pneumonia.^20,21^ In another study which considered overall cardiovascular risk, there was no difference in the likelihood of first MI or stroke after ARI by assigned cardiovascular risk.^22^ The cardiovascular complications associated with COVID-19 have prompted further research examining the impact of underlying cardiovascular risk. Generally, high prevalence of CVD risk factors have been identified in people hospitalized or dying due to COVID-19.^23^ Pooled results suggest two- to three-fold increases in the risk of severe COVID-19 outcomes among hospitalized individuals who had hypertension or diabetes.^24,25^ The degree to which age drives the associations, especially for hypertension, is unclear with variable results from age-adjusted analysis.^24–26^

There are several proposed interplaying mechanistic links between cardiovascular risk factors, ARI and CVD. Hypertension may promote immune dysregulation leading to infection, or the endothelial dysfunction caused by hypertension, hyperlipidaemia and diabetes may promote infection.^27,28^ Infectious agents, such as the SARS-CoV-2, influenza virus and *S. pneumoniae* may exacerbate atherosclerotic processes. There may be direct effects by the infectious agent of vascular cells and/or the infection may cause indirect effects through induction of haemodynamic, inflammatory and pro-coagulant processes. The release of pro-inflammatory cytokines in response to an infection can mediate atherosclerosis or directly impact plaque rupture. Endothelial dysfunction, cause by a range of cardiovascular risk factors, is a key early stage of atherosclerosis.

### Strengths and limitations

Our study’s greatest strength was the large population-based, generalizable to the UK population, linked primary and secondary care data used. We compared results across two measures of cardiovascular risk. The marked increase in MACE after ARI risk for raised QRISK2 compared with hypertension diagnosis alone is consistent with multiple cardiovascular risk factors accounted for in QRISK2.

Our selected study population should have reduced confounding; we only included individuals without chronic health conditions who are not thought to be at high-risk of ARI and ARI-related complications and not currently recommended for influenza vaccination in England. Among individuals who are eligible for influenza vaccination, health, lifestyle and behavioural differences exist between those who accept and decline vaccination.^29^ However, the selective study population did prevent comparison of ARI and MACE after ARI incidence among individuals with raised cardiovascular risk to those with established CVD.

In England, ARI diagnoses in primary and secondary care are primarily based on clinical judgement, with the majority not laboratory-confirmed. We only included codes most likely representative of systemic infection, which plausibly induce atherosclerotic processes. However, clinically-diagnosed influenza is poorly defined.^30^ Under-ascertainment of some ARIs, particularly ILI, is more likely due to short-lived illness. Conversely, due to its severity, pneumonia will most often result in healthcare attendance. The differences in influenza and pneumonia presentation and capture in EHR data may account for the divergence in ILI and pneumonia incidence among individuals with raised cardiovascular risk.

We may have misclassified QRISK2 score due to missing data for variables used in its calculation. Our similar results from recorded and calculated QRISK2 scores suggest minimal misclassification. However, this does not account for misclassification in those who attend primary care less frequently, with consultation frequency adjusted for in our analysis of ARI incidence. We used a pragmatic definition for hypertension based on coded diagnoses only; this corresponds to how in primary care, if eligible, individuals would be identified for vaccination. We likely captured both controlled and uncontrolled hypertension; from stratified analysis 38% of those with diagnosed hypertension had not been prescribed antihypertensives in the year before ARI. Conversely, only 3% of individuals with no hypertension diagnoses were prescribed antihypertensives. Possible misclassification may have biased results towards the null; this could explain the small association between hypertension and ARI compared with the larger associations for QRISK2 and, in the UK Biobank study, blood pressure reading defined hypertension.

We assumed MACE occurring after ARI was a result of infection. Yet, a proportion of the events may be unrelated, particularly those several months after infection. The possibility of unrelated events is consistent with our finding of a longer median time between ARI and MACE in individuals with raised cardiovascular risk, who are more likely to have a MACE regardless of ARI.

### Clinical and research implications

Our findings emphasize the importance of improved cardiovascular risk management, which may lessen ARI incidence and its cardiovascular consequences. The COVID-19 pandemic has highlighted this need.^31^ Cardiovascular preventive treatments, particularly among those with QRISK2 ≥10%, could reduce ARI-related complications. A meta-analysis of nine observational studies showed a 41% reduction in the odds of post-pneumonia 30-day mortality among statin users.^32^ Targeted interventions after ARI identification for individuals with QRISK2 ≥10% should be considered; a recent CPRD-based study found aspirin use among individuals hospitalized for pneumonia resulted in 54% and 30% reductions in MI and stroke, respectively.^33^

Typically, individuals with raised cardiovascular risk do not receive seasonal influenza or pneumococcal vaccines. Previous RCTs have demonstrated influenza vaccine provides secondary prevention of CVD.^7^ No RCTs or observational studies have investigated the use of vaccines for primary prevention of cardiovascular complications, such as among those with QRISK2 ≥10%. Extending influenza vaccination to those with QRISK2 ≥10% would, based on the most recent year of 2017/18 in our dataset, include approximately 150,000 individuals in England. Extending vaccination would not only reduce cardiovascular complications in those with QRISK2 ≥10%, but also lessen influenza incidence and associated complications such as hospitalizations and mortality.

Previous SCCS using national laboratory surveillance datasets linked to EHRs showed a range of respiratory viruses, including influenza virus and *S. pneumoniae*, trigger MI and stroke.^3,4^ Determining whether these organisms, along with SARS-CoV-2, lead to increases in MACE in people with raised cardiovascular risk would inform potential expansion of vaccine prioritization.

## Conclusions

In conclusion, our results suggest people with raised cardiovascular risk have an increased incidence of ARI, particularly pneumonia, and MACE after ARI. QRISK2 score provides a better measure for risk of a first cardiovascular event following ARI than hypertension diagnosis alone. Therefore, QRISK2 score could be used to not only identify individuals who require cardiovascular risk management but also targeted ARI prevention and treatment.

## Data Availability

The data used for this study were obtained from the Clinical Practice Research Datalink (CPRD). All CPRD data are available via an application to the Independent Scientific Advisory Committee (see https://www.cprd.com/Data-access). Data acquisition is associated with a fee and data protection requirements.

## Acknowledgements

This study is based on anonymized data from the Clinical Practice Research Datalink (CPRD) obtained under licence from the UK Medicines and Healthcare products Regulatory Agency. The data is provided by patients and collected by the NHS as part of their care and support. The interpretation and conclusions contained in this study are those of the authors alone.

## Contributions

C.W.-G. conceived the study. J.A.D. lead the design of the study with supervision from C.W.-G., L.S. and A.B., and further methodological contributions from H.I.M., D.G., E.H. and R.P. J.A.D lead the development and collation of code lists used to define the variables used in the study with assistance from H.I.M and H.F., and supervision from C.W.-G., L.S. and A.B. E.H. led the development of the QRISK2 coding used in the study. J.A.D. conducted the analysis with statistical input from D.G. J.A.D wrote the original manuscript draft. All authors reviewed and commented on the manuscript, and approved the final version.

## Data sharing

The data used for this study were obtained from the Clinical Practice Research Datalink (CPRD). All CPRD data are available via an application to the Independent Scientific Advisory Committee (see https://www.cprd.com/Data-access). Data acquisition is associated with a fee and data protection requirements. This manuscript is supported by code lists used to define each health condition, which have been made openly available at DOI: *awaiting*.

