## supplementary material for "Underlying cardiovascular risk and major adverse cardiovascular events after acute respiratory infection: a population-based cohort study of over 4.2 million individuals in England, 2008-2018"

**Supplementary Appendix**

### **Appendix 1. Definitions used for excluded health conditions**

| **Health condition** | **Study definition** |
| --- | --- |
| Cardiovascular disease (CVD) | Any previous clinical diagnosis, major intervention for, or clinical review specific to CVD including heart disease (congenital or otherwise), heart failure, stroke or transient ischaemic attack. |
| Chronic liver disease | Any previous clinical diagnosis of, or clinical review specific to, chronic liver disease including cirrhosis, oesophageal varices, biliary atresia and chronic hepatitis. |
| Chronic kidney disease (CKD) | Any previous clinical diagnosis of, or clinical review specific to, CKD stages 3-5, history of dialysis or renal transplant. Or with estimated glomerular filtration rate to classify CKD stages 3-5.^1^ Only stages 4-5 excluded from sensitivity analysis using pneumococcal vaccine recommendations. |
| Chronic respiratory disease (not asthma) | Any previous clinical diagnosis of, or clinical review specific to, chronic respiratory disease, including chronic obstructive pulmonary disease, emphysema, bronchitis, cystic fibrosis, or fibrosing interstitial lung diseases. |
| Asthma | Any previous clinical diagnosis of, or clinical review specific to, asthma with at least two prescriptions of inhaled steroids in the year before baseline. Or any previous hospitalisation for asthma. Not excluded from sensitivity analysis using pneumococcal vaccine recommendations. |
| Chronic neurological disease | Any previous clinical diagnosis of, or clinical review specific to, a neurological disease such as Parkinson’s disease, motor neurone disease, multiple sclerosis (MS), cerebral palsy, dementia or a learning/intellectual disability. Not excluded from sensitivity analysis using pneumococcal vaccine recommendations. |
| Diabetes mellitus | Any previous diagnosis of, or clinical review specific to, diabetes mellitus, or with a prescription for medication used to treat diabetes. Only treated diabetes excluded from sensitivity analysis using pneumococcal vaccine recommendations. |
| Asplenia/sickle cell disease | Any previous clinical diagnosis of, or clinical review specific to, asplenia or dysfunction of the spleen (including sickle cell disease but not sickle cell trait). |
| Severe obesity | Latest body mass index before baseline was ≥40 kg/m^2^. Not excluded from sensitivity analysis using pneumococcal vaccine recommendations. |
| Immunosuppression | Any previous clinical diagnosis of, or clinical review specific to, HIV, solid organ transplant or other permanent immunosuppression (such as genetic conditions compromising immune function). |
|  | Previous clinical diagnosis of, or clinical review specific to, aplastic anaemia or haematological malignancy, or receiving a bone marrow or stem cell transplant in the 2 years before baseline. |
|  | Previous clinical diagnosis of, or clinical review specific to, other/unspecified immune deficiency or receiving chemotherapy or radiotherapy in the year before baseline. |
|  | Prescription of biological therapy or at least 2 prescriptions for oral steroids or other immunosuppressants including DMARDS, Methotrexate, Azathioprine, or corticosteroid injections in the year before baseline. |

### **Appendix 2. Acute respiratory infection episode structure**

Records containing ARI codes were identified in both CPRD and HES. All ARI codes were used to define the primary outcome (ARI) and subsets were used to define the secondary outcomes of influenza/influenza-like-illness (ILI) and pneumonia.

To account for multiple consultations related to one illness, CPRD or HES records dated within 28 days of each other were regarded as part of the same illness-episode. The earliest record in the episode determined the date of illness for the primary outcome of ARI and for the secondary outcome of influenza/ILI. By comparison, for the secondary outcome of pneumonia, the earliest record which was coded as pneumonia determined the date of illness. Appendix 2 figure 1 below illustrates the episode structure. Influenza/ILI and pneumonia episodes were structured differently to account for the aetiological differences in the conditions. In an episode where there was an influenza/ILI record, but this was not the first record, it is likely that the original presentation was due to influenza/ILI but not identified as such at the time. Conversely, pneumonia can develop and worsen over time from the original infection.

In analysis of ARI outcomes, to account for multiple episodes of illness per patient, an "observation” period ended at the date of ARI and a new “observation” period begin. In analysis of MACE after ARI, an ARI episode triggered a one-year follow-up. Further ARI episodes which occurred in that further up were counted and adjusted for in analysis.

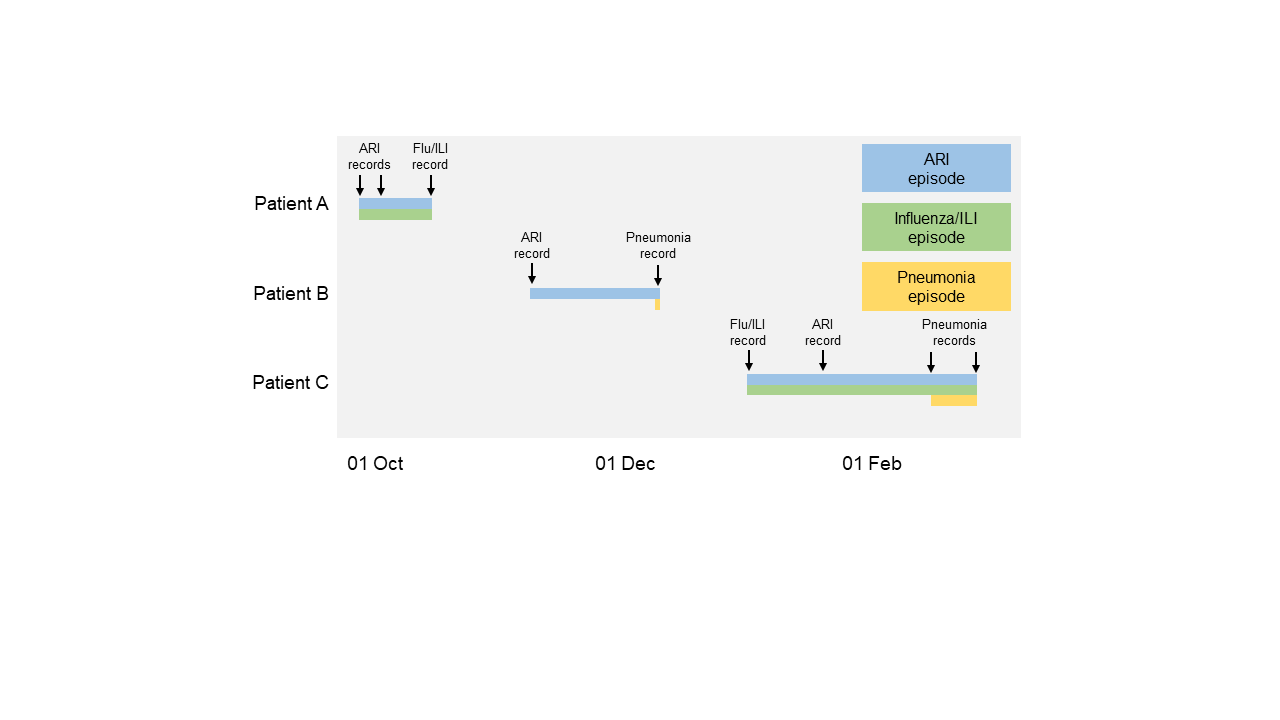

**Appendix 2 figure 1. Example patient episode classifications**

### **Appendix 3. Calculating QRISK2 scores**

An individual’s QRISK2 score is calculated based on age, sex, ethnicity, deprivation score from linked Townsend data, diabetes, family history of coronary heart disease in a first degree relative <60 years, atrial fibrillation, chronic kidney disease stage 4 or 5, rheumatoid arthritis, ratio of total serum cholesterol to high density lipoprotein cholesterol, systolic blood pressure, treated hypertension, body-mass index, and smoking status.^2^ We calculated QRISK2 scores using the algorithms supplied online (<https://qrisk.org/>) and Quality and Outcomes Framework Read and SNOMED codes^3^ (with the exception of chronic kidney disease stage 4 or 5 which used all available Read and SNOMED codes). A population-average imputation approach was used to account for missing data, to reflect the QRISK2 algorithm used in clinical practice. Our QRISK2 score process is published at <https://zenodo.org/record/3981238>.

Of note diabetes, chronic kidney disease and morbid obesity are determinants of a higher QRISK2 score, but individuals with these conditions were excluded from our study (existing eligibility for influenza vaccination).

### **Appendix 4. Comparing results from recorded and calculated QRISK2 scores**

In main analyses, our classification of cardiovascular risk level based on QRISK2 was done using our own algorithm. To validate the results obtained from our algorithm we repeated the main analyses for ARI outcome and MACE after ARI restricted to patients with a QRISK2 score recorded in CPRD directly by GPs in 2015-2017. We chose this time period as our algorithm was based on the 2015 version of QRISK2, and minimal changes were made to QRISK2 from 2015-2017.

In CPRD GOLD we identified patients with QRISK2 scores using Read codes “22W..00” , “38DF.00” or “38DP.00” from the Clinical file. If any of these three codes were recorded for consultations between 2015-2017, then the corresponding results was obtained from the Additional file. In CPRD Aurum, we used SNOMED codes “718087004”, “763244005”, “1085871000000105”, “1656451000006101”, “1656461000006104”, “810931000000108” from the Observation file.

In our subpopulation of patients with recorded QRISK2 scores we started follow-up from the latest of first recorded QRISK2 on or after 1 January 2015 or start of follow-up date from main analysis. Follow-up ended at the earliest of 31 December 2017 or end of follow-up date from main analysis.

28,579 patients had at least one QRISK2 score recorded from 2015-2017, of whom 242 had a MACE after an ARI. Similar results were obtained from recorded and calculated QRISK2 scores for both the association between raised cardiovascular risk and ARI and raised cardiovascular risk and MACE after ARI (**appendix 3 tables 1 and 2**).

**Appendix 4 Table 1.** **Crude and adjusted incidence rate ratios for the association between QRISK2 score and ARI, by QRISK2 score identification method**

| **Cardiovascular risk method** | **No. of**  **events** | **Rate per 1,000 person-years** | **Crude**  **IRR (95% CI)** | **Alcohol intake adjusted**  **IRR (95% CI)** |
| --- | --- | --- | --- | --- |
| Recorded QRISK2 ≥10% | 6,018 | 27·6 (26·8-28·4) | 1·41 (1·37-1·46) | 1·42 (1·38-1·47) |
| Recorded QRISK2 <10% | 22,561 | 19·7 (19·5-20·0) | 1 | 1 |
| Calculated QRISK2 ≥10% | 4,296 | 29·6 (28·6-30·6) | 1·48 (1·43-1·54) | 1·40 (1·35-1·46) |
| Calculated QRISK2 <10% | 24,283 | 20·0 (19·7-20·3) | 1 | 1 |

Total person-years per 1,000: calculated QRISK2 ≥10% = 145·30, calculated QRISK2 <10% = 1,215·49, recorded QRISK2 ≥10% = 218·09 and recorded QRISK2 <10% = 1,142·70.

**Appendix 4 Table 2. Crude and adjusted hazard ratios for the association between QRISK2 score and MACE after acute respiratory infection, by QRISK2 score identification method**

| **Cardiovascular risk method** | **No. of**  **events** | **Rate per 1,000 person-years** | **Crude**  **HR (95% CI)** | **Alcohol intake adjusted**  **HR (95% CI)** |
| --- | --- | --- | --- | --- |
| Recorded QRISK2 ≥10% | 110 | 22·6 (18·8-27·3) | 3·31 (2·57-4·26) | 3·34 (2·58-4·32) |
| Recorded QRISK2 <10% | 132 | 6·8 (5·7-8·1) | 1 | 1 |
| Calculated QRISK2 ≥10% | 89 | 25·3 (20·6-31·2) | 3·43 (2·65-4·46) | 3·39 (2·60-4·42) |
| Calculated QRISK2 <10% | 153 | 7·4 (6·3-8·6) | 1 | 1 |

Total person-years per 1,000: calculated QRISK2 ≥10% = 3·51, calculated QRISK2 <10% = 20·75, recorded QRISK2 ≥10% = 4·87 and recorded QRISK2 <10% = 19·40.

### **Table S1. Baseline demographic and lifestyle characteristics of included study population separated by database**

|  | **Gold**  **n = 773,362** | **Aurum**  **n = 3,439,568** |
| --- | --- | --- |
| Age (years) |  |  |
| 40-44 | 316,376 (40·9%) | 1,603,993 (46·6%) |
| 45-49 | 152,207 (19·7%) | 630,690 (18·3%) |
| 50-54 | 121,385 (15·7%) | 490,817 (14·3%) |
| 55-59 | 99,557 (12·9%) | 391,062 (11·4%) |
| 60-64 | 83,837 (10·8%) | 323,006 (9·4%) |
| Sex | n = 773,351 | n = 3,439,547 |
| Male | 405,608 (52·4%) | 1,820,953 (52·9%) |
| Female | 367,743 (47·6%) | 1,618,594 (47·1%) |
| Ethnicity | n = 642,421 | n = 3,060,297 |
| White | 580,960 (90·4%) | 2,661,147 (87·0%) |
| South Asian | 24,419 (3·8%) | 170,512 (5·6%) |
| Black | 18,343 (2·9%) | 135,927 (4·4%) |
| Mixed/Other | 18,699 (2·9%) | 92,711 (3·0%) |
| Townsend quintile | n = 772,985 | n = 3,434,620 |
| 1 (least deprived) | 190,835 (24·7%) | 813,835 (23·7%) |
| 2 | 180,189 (23·3%) | 725,502 (21·1%) |
| 3 | 158,379 (20·5%) | 667,300 (19·4%) |
| 4 | 138,537 (17·9%) | 600,650 (17·5%) |
| 5 (most deprived) | 105,045 (13·6%) | 627,333 (18·3%) |
| BMI category* | n = 772,985 | n = 2,803,225 |
| Underweight (<18·5 kg/m2) | 9,269 (1·4%) | 41,733 (1·5%) |
| Normal weight (18·5-24·9 kg/m2) | 271,221 (42·1%) | 1,178,462 (42·0%) |
| Overweight (25·0-29·9 kg/m2) | 238,478 (37·0%) | 1,036,223 (37·0%) |
| Obese (30·0-39·9 kg/m2) | 125,411 (19·5%) | 546,807 (19·5%) |
| Smoking status* | n = 747,925 | n = 3,334,866 |
| Non-smoker | 354,909 (47·5%) | 1,332,010 (39·9%) |
| Current smoker | 193,384 (25·9%) | 883,323 (26·5%) |
| Ex-smoker | 199,632 (26·7%) | 1,119,533 (33·6%) |
| Alcohol consumption* | n = 678,452 | n = 2,996,042 |
| Not a heavy drinker | 654,536 (96·5%) | 2,822,800 (94·2%) |
| Heavy drinker | 26,109 (3·5%) | 173,242 (5·8%) |

Data are n (%). *Closest measure before start of follow-up.

### **Table S2.** **Database comparison of acute respiratory infection incidence rates and incidence rate ratios**

| **Outcome** | **Cardiovascular risk** | **Database** | **Rate per 1,000 person-years**  **(95% CI)** | | **Crude**  **IRR (95% CI)** | **Age and sex-adjusted IRR (95% CI)** | **Fully-adjusted* IRR (95% CI)** |
| --- | --- | --- | --- | --- | --- | --- | --- |
|  |  |  | **High risk** | **Low risk** |  |  |  |
| ARI | Hypertension | GOLD | 39·5 (38·7-40·4) | 29·9 (29·6-30·1) | 1·32 (1·29-1·35) | 1·29 (1·26-1·32) | 1·08 (1·05-1·11) |
|  |  | Aurum | 40·5 (40·1-40·9) | 29·0 (28·9-29·2) | 1·39 (1·37-1·40) | 1·34 (1·32-1·35) | 1·05 (1·03-1·06) |
|  |  | GOLD and Aurum | 40·3 (40·0-40·7) | 29·1 (29·0-29·2) | 1·38 (1·36-1·39) | 1·33 (1·32-1·34) | 1·04 (1·03-1·05) |
|  |  | Meta-analysis of GOLD and Aurum | - | - | 1·36 (1·29-1·43) | 1·32 (1·27-1·37) | 1·06 (1·03-1·09) |
|  | QRISK2 ≥10% | GOLD | 43·9 (42·9-44·8) | 29·6 (29·3-29·8) | 1·49 (1·45-1·52) | - | 1·37 (1·34-1·41) |
|  |  | Aurum | 43·8 (43·3-44·2) | 28·6 (28·5-28·8) | 1·52 (1·51-1·54) | - | 1·39 (1·37-1·40) |
|  |  | GOLD and Aurum | 43·8 (43·4-44·2) | 28·8 (28·7-28·9) | 1·52 (1·50-1·53) | - | 1·39 (1·37-1·40) |
|  |  | Meta-analysis of GOLD and Aurum | - | - | 1·51 (1·48-1·54) | - | 1·39 (1·37-1·40) |
| Influenza / ILI | Hypertension | GOLD | 6·2 (5·9-6·5) | 5·6 (5·5-5·7) | 1·10 (1·05-1·16) | 1·22 (1·16-1·29) | 1·01 (0·96-1·07) |
|  |  | Aurum | 6·3 (6·1-6·4) | 5·5 (5·4-5·5) | 1·14 (1·12-1·17) | 1·25 (1·22-1·28) | 0·98 (0·95-1·00) |
|  |  | GOLD and Aurum | 6·3 (6·1-6·4) | 5·5 (5·4-5·5) | 1·14 (1·11-1·16) | 1·25 (1·22-1·27) | 0·98 (0·96-1·00) |
|  |  | Meta-analysis of GOLD and Aurum | - | - | 1·13 (1·09-1·16) | 1·25 (1·22-1·27) | 0·99 (0·96-1·01) |
|  | QRISK2 ≥10% | GOLD | 5·1 (4·8-5·3) | 5·7 (5·6-5·8) | 0·89 (0·84-0·94) | - | 0·83 (0·78-0·88) |
|  |  | Aurum | 5·4 (5·3-5·5) | 5·5 (5·5-5·6) | 0·97 (0·95-0·99) | - | 0·89 (0·87-0·91) |
|  |  | GOLD and Aurum | 5·4 (5·3-5·5) | 5·6 (5·5-5·6) | 0·96 (0·94-0·98) | - | 0·88 (0·86-0·90) |
|  |  | Meta-analysis of GOLD and Aurum | - | - | 0·93 (0·86-1·01) | - | 0·87 (0·81-0·92) |
| Pneumonia | Hypertension | GOLD | 1·9 (1·7-2·1) | 1·3 (1·3-1·4) | 1·46 (1·32-1·61) | 1·25 (1·13-1·39) | 1·07 (0·97-1·19) |
|  |  | Aurum | 2·4 (2·3-2·5) | 1·6 (1·5-1·6) | 1·61 (1·55-1·68) | 1·33 (1·28-1·39) | 1·12 (1·07-1·17) |
|  |  | GOLD and Aurum | 2·3 (2·2-2·4) | 1·5 (1·5-1·5) | 1·59 (1·53-1·65) | 1·32 (1·27-1·38) | 1·12 (1·07-1·16) |
|  |  | Meta-analysis of GOLD and Aurum | - | - | 1·55 (1·40-1·69) | 1·31 (1·25-1·38) | 1·11 (1·07-1·16) |
|  | QRISK2 ≥10% | GOLD | 3·1 (2·9-3·3) | 1·2 (1·2-1·3) | 2·62 (2·41-2·86) | - | 2·37 (2·17-2·60) |
|  |  | Aurum | 3·5 (3·4-3·6) | 1·4 (1·4-1·5) | 2·59 (2·50-2·69) | - | 2·31 (2·23-2·39) |
|  |  | GOLD and Aurum | 3·5 (3·4-3·6) | 1·4 (1·4-1·4) | 2·60 (2·52-2·69) | - | 2·32 (2·25-2·40) |
|  |  | Meta-analysis of GOLD and Aurum | - | - | 2·60 (2·51-2·68) | - | 2·32 (2·24-2·39) |

*Hypertension models adjusted for: age, sex, ethnicity, socioeconomic status, BMI, alcohol intake, smoking status and consultation frequency. QRISK2 models adjusted for: alcohol intake and consultation frequency.

### **Table S3. Crude and adjusted incidence rate ratios for the association between cardiovascular risk and ARI among sensitivity analysis study population**

| **Outcome** | **Cardiovascular risk** | **No. of**  **events** | **Rate per 1,000 person-years (95% CI)** | **Crude**  **IRR (95% CI)** | **Age and sex-adjusted**  **IRR (95% CI)** | **Fully-adjusted***  **IRR (95% CI)** |
| --- | --- | --- | --- | --- | --- | --- |
| Acute respiratory infection | Hypertension | 96,203 | 42·7 (42·4-43·1) | 1·40 (1·38-1·41) | 1·34 (1·33-1·36) | 1·04 (1·03-1·05) |
|  | No hypertension | 558,302 | 30·4 (30·3-30·5) | 1 | 1 | 1 |
|  | QRISK2 ≥10% | 96,193 | 45·8 (45·4-46·2) | 1·51 (1·50-1·52) | - | 1·37 (1·36-1·39) |
|  | QRISK2 <10% | 558,312 | 30·1 (30·0-30·2) | 1 | - | 1 |
| Influenza/ILI | Hypertension | 14,436 | 6·4 (6·3-6·5) | 1·14 (1·12-1·17) | 1·25 (1·23-1·27) | 0·98 (0·96-1·00) |
|  | No hypertension | 102,207 | 5·6 (5·5-5·6) | 1 | 1 | 1 |
|  | QRISK2 ≥10% | 11,573 | 5·5 (5·4-5·6) | 0·97 (0·95-0·99) | - | 0·88 (0·86-0·90) |
|  | QRISK2 <10% | 105,070 | 5·7 (5·6-5·7) | 1 | - | 1 |
| Pneumonia | Hypertension | 5,700 | 2·5 (2·5-2·6) | 1·61 (1·56-1·67) | 1·35 (1·30-1·39) | 1·11 (1·07-1·16) |
|  | No hypertension | 29,987 | 1·6 (1·6-1·7) | 1 | 1 | 1 |
|  | QRISK2 ≥10% | 7,798 | 3·7 (3·6-3·8) | 2·59 (2·51-2·67) | - | 2·30 (2·23-2·37) |
|  | QRISK2 <10% | 27,889 | 1·5 (1·5-1·5) | 1 | - | 1 |

Total person-years per 1,000: hypertension = 2,251·4, no hypertension = 18,373·1, QRISK2 ≥10% = 2,099·5 and QRISK2 <10% = 18,524·0. LRT p-values all <0·001. *Hypertension models adjusted for: age, sex, ethnicity, socioeconomic status, BMI, alcohol intake, smoking status and consultation frequency. QRISK2 models adjusted for: alcohol intake and consultation frequency.

### **Table S4. MACE after influenza/ILI incidence rates and hazard ratios by cardiovascular risk group**

| **Outcome** | **Cardiovascular risk** | **No. of events** | **Rate per 1,000 person-years (95% CI)** | **Crude**  **HR (95% CI)** | **Age and sex-adjusted**  **HR (95% CI)** | **Fully-adjusted***  **HR (95% CI)** |
| --- | --- | --- | --- | --- | --- | --- |
| Any MACE | Hypertension | 99 | 8·0 (6·6-9·8) | 2·24 (1·79-2·80) | 2·13 (1·70-2·66) | 2·07 (1·60-2·67) |
|  | No hypertension | 352 | 3·7 (3·3-4·1) | 1 | 1 | 1 |
|  | QRISK2 ≥10% | 116 | 11·5 (9·7-13·9) | 3·36 (2·72-4·15) | - | 3·35 (2·70-4·17) |
|  | QRISK2 <10% | 335 | 3·4 (3·1-3·8) | 1 | - | 1 |
| ACS | Hypertension | 34 | 2·8 (2·0-3·9) | 1·99 (1·36-2·90) | 1·87 (1·28-2·73) | 1·97 (1·30-2·99) |
|  | No hypertension | 136 | 1·4 (1·2-1·7) | 1 | 1 | 1 |
|  | QRISK2 ≥10% | 51 | 5·1 (3·9-6·8) | 4·15 (2·99-5·77) | - | 4·04 (2·89-5·63) |
|  | QRISK2 <10% | 119 | 1·2 (1·0-1·5) | 1 | - | 1 |
| Heart failure | Hypertension | 21 | 1·7 (1·1-2·7) | 2·85 (1·73-4·69) | 2·69 (1·63-4·43) | 2·82 (1·53-5·21) |
|  | No hypertension | 59 | 0·6 (0·5-0·8) | 1 | 1 | 1 |
|  | QRISK2 ≥10% | 22 | 2·2 (1·5-3·4) | 3·68 (2·25-6·01) | - | 3·72 (2·23-6·19) |
|  | QRISK2 <10% | 58 | 0·6 (0·5-0·8) | 1 | - | 1 |
| Stroke or TIA | Hypertension | 45 | 3·6 (2·7-5·0) | 2·33 (1·67-3·25) | 2·24 (1·60-3·13) | 2·04 (1·39-2·98) |
|  | No hypertension | 153 | 1·6 (1·4-1·9) | 1 | 1 | 1 |
|  | QRISK2 ≥10% | 41 | 4·1 (3·0-5·6) | 2·53 (1·80-3·57) | - | 2·49 (1·74-3·57) |
|  | QRISK2 <10% | 157 | 1·6 (1·4-1·9) | 1 | - | 1 |
| CVD death | Hypertension | 12 | 1·0 (0·6-1·8) | 2·81 (1·46-5·41) | 2·66 (1·38-5·12) | 3·31 (1·51-7·25) |
|  | No hypertension | 34 | 0·4 (0·3-0·5) | 1 | 1 | 1 |
|  | QRISK2 ≥10% | 13 | 1·3 (0·8-2·4) | 3·82 (2·01-7·26) | - | 4·19 (2·17-8·07) |
|  | QRISK2 <10% | 33 | 0·3 (0·2-0·5) | 1 | - | 1 |

Acute limb ischaemia not included as secondary outcome due to event numbers <10. Total person-years per 1,000: hypertension = 12·3, no hypertension = 95·7, QRISK2 ≥10% = 10·0 and QRISK2 <10% = 98·0. LRT p-values all <0·001. *Hypertension models adjusted for: age, sex, ethnicity, socioeconomic status, BMI, alcohol intake and smoking status. QRISK2 models adjusted for: alcohol intake. †Results subdivided into MI and unstable angina separately: MI fully-adjusted HR in hypertension model 2·34 (95% CI 1·48-3·71) and in QRISK2 model 4·84 (95% CI 3·36-6·97), angina fully-adjusted HR in hypertension model 0·89 (95% CI 0·30-2·61) and in QRISK2 model 2·17 (95% CI 0·96-4·91). ‡Results subdivided into stroke and TIA separately: stroke fully-adjusted HR in hypertension model 2·40 (95% CI 1·54-3·74) and in QRISK2 model 2·69 (95% CI 1·76-4·12), TIA fully-adjusted HR in hypertension model 1·49 (95% CI 0·76-2·91) and in QRISK2 model 2·47 (95% CI 1·33-4·56).

### **Table S5. MACE after pneumonia incidence rates and incidence rate ratios by cardiovascular risk group**

| **Outcome** | **Cardiovascular risk** | **No. of events** | **Rate per 1,000 person-years (95% CI)** | **Crude**  **HR (95% CI)** | **Age and sex-adjusted**  **HR (95% CI)** | **Fully-adjusted***  **HR (95% CI)** |
| --- | --- | --- | --- | --- | --- | --- |
| Any MACE | Hypertension | 341 | 105·6 (94·5-118·3) | 1·62 (1·44-1·82) | 1·58 (1·41-1·78) | 1·65 (1·44-1·89) |
|  | No hypertension | 1,265 | 63·6 (60·1-67·4) | 1 | 1 | 1 |
|  | QRISK2 ≥10% | 574 | 130·9 (120·1-142·9) | 2·17 (1·96-2·40) | - | 2·13 (1·92-2·37) |
|  | QRISK2 <10% | 1,032 | 55·1 (51·8-58·7) | 1 | - | 1 |
| ACS | Hypertension | 117 | 36·1 (30·1-43·6) | 1·73 (1·41-2·12) | 1·68 (1·37-2·06) | 1·91 (1·50-2·44) |
|  | No hypertension | 405 | 20·3 (18·4-22·5) | 1 | 1 | 1 |
|  | QRISK2 ≥10% | 217 | 49·5 (43·3-56·9) | 2·75 (2·31-3·26) | - | 2·82 (2·35-3·38) |
|  | QRISK2 <10% | 305 | 16·2 (14·5-18·2) | 1 | - | 1 |
| Heart failure | Hypertension | 145 | 44·8 (38·1-53·1) | 1·59 (1·32-1·90) | 1·56 (1·30-1·87) | 1·70 (1·37-2·11) |
|  | No hypertension | 545 | 27·4 (25·2-29·8) | 1 | 1 | 1 |
|  | QRISK2 ≥10% | 253 | 57·7 (50·9-65·9) | 2·26 (1·94-2·64) | - | 2·20 (1·87-2·58) |
|  | QRISK2 <10% | 437 | 23·3 (21·2-25·7) | 1 | - | 1 |
| Acute limb ischaemia | Hypertension | 10 | 3·1 (1·7-6·3) | 2·32 (1·12-4·80) | 2·30 (1·11-4·76) | 3·19 (1·34-7·55) |
|  | No hypertension | 25 | 1·3 (0·9-1·9) | 1 | 1 | 1 |
|  | QRISK2 ≥10% | 15 | 3·4 (2·1-6·0) | 3·21 (1·65-6·26) | - | 3·26 (1·65-6·42) |
|  | QRISK2 <10% | 20 | 1·1 (0·7-1·7) | 1 | - | 1 |
| Stroke or TIA | Hypertension | 124 | 38·4 (32·2-46·1) | 1·85 (1·52-2·26) | 1·83 (1·50-2·23) | 1·79 (1·43-2·24) |
|  | No hypertension | 400 | 20·1 (18·2-22·2) | 1 | 1 | 1 |
|  | QRISK2 ≥10% | 160 | 36·5 (31·2-42·9) | 1·71 (1·42-2·06) | - | 1·67 (1·38-2·04) |
|  | QRISK2 <10% | 364 | 19·4 (17·5-21·6) | 1 | - | 1 |
| CVD death | Hypertension | 58 | 17·9 (13·9-23·5) | 1·60 (1·20-2·14) | 1·56 (1·17-2·08) | 1·57 (1·11-2·23) |
|  | No hypertension | 219 | 11·0 (9·6-12·6) | 1 | 1 | 1 |
|  | QRISK2 ≥10% | 120 | 27·4 (22·9-33·0) | 3·02 (2·39-3·83) | - | 3·11 (2·39-4·04) |
|  | QRISK2 <10% | 157 | 8·4 (7·2-9·8) | 1 | - | 1 |

Total person-years per 1,000: hypertension = 3·2, no hypertension = 19·9, QRISK2 ≥10% = 4·4 and QRISK2 <10% = 18·7. LRT p-values all <0·001. *Hypertension models adjusted for: age, sex, ethnicity, socioeconomic status, BMI, alcohol intake and smoking status. QRISK2 models adjusted for: alcohol intake. †Results subdivided into MI and unstable angina separately: MI fully-adjusted HR in hypertension model 1·94 (95% CI 1·48-2·54) and in QRISK2 model 3·04 (95% CI 2·49-3·70), angina fully-adjusted HR in hypertension model 2·42 (95% CI 1·38-4·22) and in QRISK2 model 2·03 (95% CI 1·24-3·33). ‡Results subdivided into stroke and TIA separately: stroke fully-adjusted HR in hypertension model 1·78 (95% CI 1·42-2·23) and in QRISK2 model 1·77 (95% CI 1·45-2·15), TIA fully-adjusted HR in hypertension model 1·74 (95% CI 0·74-4·08) and in QRISK2 model 1·14 (95% CI 0·56-2·34).

### **Table S6. MACE after ARI by anti-hypertensives, statins and antiplatelets prescription status**

| **Prescribed treatment of interest** | **Cardiovascular risk** | **No. of events** | **Total person-years per 1,000** | **Rate per 1,000 person-years (95% CI)** | **Crude**  **HR (95% CI)** | **Age and sex-adjusted**  **HR (95% CI)** | **Fully-adjusted***  **HR (95% CI)** |
| --- | --- | --- | --- | --- | --- | --- | --- |
| Anti-hypertensives  (n=57,800) | Hypertension (n=42,888) | 532 | 55·6 | 9·6 (8·8-10·4) | 1·00 (0·84-1·18) | 0·86 (0·72-1·02) | 1·03 (0·83-1·27) |
|  | No hypertension (n=14,912) | 183 | 19·0 | 9·6 (8·4-11·2) | 1 | 1 | 1 |
|  | QRISK2 ≥10% (n=22,805) | 436 | 28·9 | 15·1 (13·7-16·6) | 2·45 (2·11-2·85) | - | 2·47 (2·12-2·89) |
|  | QRISK2 <10% (n=34,995) | 279 | 45·6 | 6·1 (5·4-6·9) | 1 | - | 1 |
| No anti-hypertensives  (n=469,000) | Hypertension (n=25,843) | 453 | 32·2 | 14·1 (12·8-15·4) | 2·62 (2·37-2·89) | 2·58 (2·34-2·85) | 2·56 (2·30-2·86) |
|  | No hypertension (n=443,157) | 3,001 | 534·5 | 5·6 (5·4-5·8) | 1 | 1 | 1 |
|  | QRISK2 ≥10% (n=49,332) | 1,090 | 58·1 | 18·8 (17·7-19·9) | 4·04 (3·76-4·34) | - | 3·93 (3·65-4·24) |
|  | QRISK2 <10% (n=419,668) | 2,364 | 508·7 | 4·6 (4·5-4·8) | 1 | - | 1 |
| Statins  (n=33,837) | Hypertension (n=16,865) | 246 | 21·9 | 11·3 (9·9-12·8) | 1·09 (0·91-1·30) | 1·06 (0·88-1·26) | 1·11 (0·91-1·35) |
|  | No hypertension (n=16,972) | 228 | 22·1 | 10·3 (9·1-11·8) | 1 | 1 | 1 |
|  | QRISK2 ≥10% (n=16,746) | 322 | 21·5 | 15·0 (13·4-16·7) | 2·22 (1·83-2·69) | - | 2·16 (1·77-2·63) |
|  | QRISK2 <10% (n=17,091) | 152 | 22·5 | 6·8 (5·8-8·0) | 1 | - | 1 |
| No statins  (n=492,963) | Hypertension (n=51,866) | 739 | 66·0 | 11·2 (10·4-12·1) | 2·14 (1·98-2·32) | 2·09 (1·93-2·27) | 2·13 (1·94-2·34) |
|  | No hypertension (n=441,097) | 2,956 | 531·4 | 5·6 (5·4-5·8) | 1 | 1 | 1 |
|  | QRISK2 ≥10% (n=55,391) | 1,204 | 65·5 | 18·4 (17·4-19·5) | 3·93 (3·67-4·21) | - | 3·83 (3·57-4·11) |
|  | QRISK2 <10% (n=437,572) | 2,491 | 531·8 | 4·7 (4·5-4·9) | 1 | - | 1 |
| Antiplatelets  (n=7,670) | Hypertension (n=3,658) | 88 | 4·4 | 20·0 (16·3-24·9) | 0·86 (0·65-1·13) | 0·78 (0·59-1·04) | 0·94 (0·68-1·28) |
|  | No hypertension (n=4,012) | 112 | 4·9 | 22·9 (19·1-27·8) | 1 | 1 | 1 |
|  | QRISK2 ≥10% (n=3,970) | 131 | 4·7 | 28·2 (23·7-33·7) | 1·82 (1·36-2·44) | - | 1·72 (1·27-2·33) |
|  | QRISK2 <10% (n=3,700) | 69 | 4·6 | 14·9 (11·8-19·1) | 1 | - | 1 |
| No antiplatelets  (n=519,130) | Hypertension (n=65,073) | 897 | 83·4 | 10·8 (10·1-11·5) | 2·05 (1·91-2·21) | 1·96 (1·82-2·11) | 1·97 (1·81-2·15) |
|  | No hypertension (n=454,057) | 3,072 | 548·6 | 5·6 (54-5·8) | 1 | 1 | 1 |
|  | QRISK2 ≥10% (n=68,167) | 1,395 | 82·4 | 16·9 (16·1-17·9) | 3·68 (3·45-3·93) | - | 3·60 (3·36-3·85) |
|  | QRISK2 <10% (n=450,963) | 2,574 | 549·7 | 4·7 (4·5-4·9) | 1 | - | 1 |

*Hypertension models adjusted for: age, sex, ethnicity, socioeconomic status, BMI, alcohol intake and smoking status. QRISK2 models adjusted for: alcohol intake.

### **Table S7. Database comparison of MACE after ARI incidence rates and hazard ratios by cardiovascular risk group**

| **Outcome** | **Cardiovascular risk** | **Database** | **Rate per 1,000 person-years**  **(95% CI)** | | **Crude**  **HR (95% CI)** | **Age and sex-adjusted**  **HR (95% CI)** | **Fully-adjusted* HR (95% CI)** |
| --- | --- | --- | --- | --- | --- | --- | --- |
|  |  |  | **High risk** | **Low risk** |  |  |  |
| Any MACE | Hypertension | GOLD | 11·5 (9·7-13·6) | 5·9 (5·4-6·4) | 2·03 (1·68-2·46) | 1·92 (1·59-2·33) | 2·06 (1·67-2·54) |
|  |  | Aurum | 11·2 (10·4-12·0) | 5·7 (5·5-6·0) | 2·08 (1·93-2·25) | 1·98 (1·84-2·14) | 1·97 (1·80-2·16) |
|  |  | GOLD and Aurum | 11·2 (10·5-12·0) | 5·8 (5·5-6·0) | 2·08 (1·93-2·23) | 1·97 (1·84-2·12) | 1·98 (1·83-2·15) |
|  |  | Meta-analysis of GOLD and Aurum | - | - | 2·07 (1·93-2·22) | 1·97 (1·83-2·11) | 1·98 (1·82-2·15) |
|  | QRISK2 | GOLD | 17·2 (15·0-19·9) | 5·1 (4·6-5·6) | 3·37 (2·85-4·00) | - | 3·35 (2·81-3·99) |
|  |  | Aurum | 17·6 (16·7-18·6) | 4·7 (4·5-4·9) | 3·80 (3·55-4·07) | - | 3·70 (3·44-3·97) |
|  |  | GOLD and Aurum | 17·5 (16·7-18·5) | 4·8 (4·6-5·0) | 3·65 (3·42-3·89) | - | 3·65 (3·42-3·89) |
|  |  | Meta-analysis of GOLD and Aurum | - | - | 3·67 (3·27-4·06) | - | 3·63 (3·35-3·91) |
| ACS | Hypertension | GOLD | 4·8 (3·7-6·2) | 2·2 (1·9-2·6) | 2·24 (1·66-3·02) | 2·10 (1·55-2·83) | 2·18 (1·54-3·08) |
|  |  | Aurum | 4·1 (3·7-4·6) | 2·0 (1·9-2·2) | 2·18 (1·92-2·48) | 2·06 (1·81-2·34) | 2·12 (1·83-2·46) |
|  |  | GOLD and Aurum | 4·2 (3·8-4·7) | 2·1 (1·9-2·2) | 2·19 (1·95-2·46) | 2·06 (1·83-2·32) | 2·13 (1·86-2·44) |
|  |  | Meta-analysis of GOLD and Aurum | - | - | 2·19 (1·93-2·45) | 2·07 (1·82-2·31) | 2·13 (1·84-2·42) |
|  | QRISK2 | GOLD | 7·0 (5·7-8·8) | 1·9 (1·6-2·2) | 3·66 (2·80-4·79) | - | 3·79 (2·87-5·00) |
|  |  | Aurum | 7·0 (6·5-7·7) | 1·6 (1·5-1·7) | 4·56 (4·08-5·10) | - | 4·48 (3·99-5·03) |
|  |  | GOLD and Aurum | 7·0 (6·5-7·6) | 1·6 (1·5-1·7) | 4·42 (3·98-4·89) | - | 4·37 (3·93-4·86) |
|  |  | Meta-analysis of GOLD and Aurum | - | - | 4·22 (3·36-5·01) | - | 4·30 (3·70-4·89) |
| Heart failure | Hypertension | GOLD | 4·2 (3·2-5·6) | 1·7 (1·5-2·0) | 2·53 (1·83-3·49) | 2·39 (1·74-3·30) | 2·83 (1·96-4·08) |
|  |  | Aurum | 3·2 (2·8-3·6) | 1·7 (1·6-1·9) | 1·96 (1·69-2·26) | 1·85 (1·60-2·13) | 1·96 (1·65-2·32) |
|  |  | GOLD and Aurum | 3·3 (2·9-3·7) | 1·7 (1·6-1·9) | 2·04 (1·79-2·32) | 1·92 (1·69-2·19) | 2·08 (1·79-2·42) |
|  |  | Meta-analysis of GOLD and Aurum | - | - | 2·11 (1·62-2·59) | 1·99 (1·53-2·45) | 2·24 (1·44-3·04) |
|  | QRISK2 | GOLD | 5·5 (4·3-7·1) | 1·6 (1·3-1·9) | 3·50 (2·58-4·73) | - | 3·43 (2·51-4·70) |
|  |  | Aurum | 5·5 (5·0-6·1) | 1·4 (1·3-1·5) | 4·10 (3·62-4·63) | - | 3·93 (3·45-4·48) |
|  |  | GOLD and Aurum | 5·5 (5·0-6·0) | 1·4 (1·3-1·5) | 4·00 (3·57-4·49) | - | 3·85 (3·42-4·34) |
|  |  | Meta-analysis of GOLD and Aurum | - | - | 3·99 (3·53-4·45) | - | 3·84 (3·37-4·31) |
| Acute limb ischaemia | Hypertension | GOLD | 0·0 (0·0-0·0) | 0·1 (0·1-0·2) | - | - | - |
|  |  | Aurum | 0·3 (0·2-0·5) | 0·1 (0·1-0·1) | 3·49 (2·15-5·68) | 3·32 (2·03-5·43) | 5·72 (3·25-10·08) |
|  |  | GOLD and Aurum | 0·3 (0·2-0·4) | 0·1 (0·1-0·1) | 2·98 (1·85-4·78) | 2·82 (1·74-4·55) | 4·63 (2·68-7·99) |
|  |  | Meta-analysis of GOLD and Aurum | - | - | - | - | - |
|  | QRISK2 | GOLD | 0·3 (0·1-1·3) | 0·1 (0·0-0·2) | 4·18 (0·99-17·68) | - | 4·21 (0·99-17·85) |
|  |  | Aurum | 0·5 (0·4-0·7) | 0·1 (0·0-0·1) | 7·56 (4·76-12·00) | - | 7·34 (4·56-11·80) |
|  |  | GOLD and Aurum | 0·5 (0·4-0·7) | 0·1 (0·1-0·1) | 7·15 (4·62-11·07) | - | 6·93 (4·43-10·83) |
|  |  | Meta-analysis of GOLD and Aurum | - | - | 7·03 (3·70-10·35) | - | 6·85 (3·53-10·18) |
| Stroke or TIA | Hypertension | GOLD | 4·0 (3·1-5·4) | 2·0 (1·7-2·4) | 2·06 (1·50-2·85) | 1·96 (1·42-2·71) | 2·12 (1·50-2·99) |
|  |  | Aurum | 4·1 (3·7-4·6) | 2·0 (1·9-2·2) | 2·17 (1·91-2·46) | 2·10 (1·84-2·38) | 1·99 (1·72-2·30) |
|  |  | GOLD and Aurum | 4·1 (3·7-4·6) | 2·0 (1·9-2·1) | 2·15 (1·91-2·42) | 2·08 (1·84-2·34) | 2·01 (1·75-2·29) |
|  |  | Meta-analysis of GOLD and Aurum | - | - | 2·15 (1·90-2·41) | 2·08 (1·83-2·33) | 2·01 (1·74-2·28) |
|  | QRISK2 | GOLD | 5·6 (4·4-7·2) | 1·8 (1·5-2·1) | 3·08 (2·29-4·14) | - | 2·91 (2·14-3·95) |
|  |  | Aurum | 5·3 (4·9-5·9) | 1·8 (1·7-2·0) | 2·98 (2·65-3·35) | - | 2·93 (2·59-3·31) |
|  |  | GOLD and Aurum | 5·4 (4·9-5·9) | 1·8 (1·7-1·9) | 2·99 (2·68-3·34) | - | 2·93 (2·62-3·28) |
|  |  | Meta-analysis of GOLD and Aurum | - | - | 2·99 (2·67-3·32) | - | 2·93 (2·59-3·26) |
| CVD death | Hypertension | GOLD | 1·4 (0·9-2·3) | 0·7 (0·5-0·9) | 2·06 (1·19-3·56) | 1·96 (1·13-3·40) | 2·31 (1·13-4·71) |
|  |  | Aurum | 1·5 (1·2-1·8) | 0·8 (0·7-0·8) | 2·12 (1·72-2·63) | 2·00 (1·62-2·47) | 2·13 (1·65-2·74) |
|  |  | GOLD and Aurum | 1·5 (1·2-1·8) | 0·7 (0·7-0·8) | 2·11 (1·73-2·58) | 1·99 (1·63-2·43) | 2·15 (1·69-2·73) |
|  |  | Meta-analysis of GOLD and Aurum | - | - | 2·11 (1·69-2·54) | 2·00 (1·60-2·39) | 2·15 (1·62-2·67) |
|  | QRISK2 | GOLD | 2·5 (1·8-3·7) | 0·5 (0·4-0·7) | 4·58 (2·87-7·31) | - | 4·86 (2·91-8·11) |
|  |  | Aurum | 2·7 (2·3-3·1) | 0·6 (0·5-0·6) | 4·80 (4·00-5·76) | - | 4·80 (3·92-5·87) |
|  |  | GOLD and Aurum | 2·6 (2·3-3·0) | 0·6 (0·5-0·6) | 4·77 (4·03-5·66) | - | 4·81 (3·99-5·81) |
|  |  | Meta-analysis of GOLD and Aurum | - | - | 4·77 (3·95-5·58) | - | 4·81 (3·89-5·72) |

In meta-analysis the between database heterogeneity was assessed using the I^2^ statistic. The I^2^ for all pooled estimates was 0, with the exception of; QRISK2 and MACE (crude I^2^=44%, fully-adjusted I^2^=11%), QRISK2 and ACS (crude I^2^=60%, fully-adjusted I^2^=23%), and hypertension and heart failure (crude and age- and sex-adjusted I^2^=38%, fully-adjusted I^2^=58%). *Hypertension models adjusted for: age, sex, ethnicity, socioeconomic status, BMI, alcohol intake and smoking status. QRISK2 models adjusted for: alcohol intake.

### **Table S8. Crude and adjusted hazard ratios for the association between cardiovascular risk and MACE after ARI among sensitivity analysis study population**

| **Outcome** | **Cardiovascular risk** | **No. of events** | **Rate per 1,000 person-years (95% CI)** | **Crude**  **HR (95% CI)** | **Age and sex-adjusted**  **HR (95% CI)** | **Fully-adjusted***  **HR (95% CI)** |
| --- | --- | --- | --- | --- | --- | --- |
| Any MACE | Hypertension | 1,263 | 11·6 (11·0-12·3) | 2·12 (1·98-2·26) | 2·04 (1·91-2·18) | 2·04 (1·89-2·19) |
|  | No hypertension | 3,560 | 5·9 (5·7-6·1) | 1 | 1 | 1 |
|  | QRISK2 ≥10% | 1,820 | 17·8 (17·0-18·6) | 3·70 (3·49-3·92) | - | 3·61 (3·40-3·84) |
|  | QRISK2 <10% | 3,003 | 4·9 (4·7-5·1) | 1 | - | 1 |
| ACS | Hypertension | 437 | 4·0 (3·7-4·4) | 2·10 (1·88-2·34) | 2·01 (1·80-2·24) | 2·08 (1·83-2·36) |
|  | No hypertension | 1,240 | 2·0 (1·9-2·2) | 1 | 1 | 1 |
|  | QRISK2 ≥10% | 695 | 6·8 (6·3-7·3) | 4·31 (3·92-4·75) | - | 4·29 (3·88-4·74) |
|  | QRISK2 <10% | 982 | 1·6 (1·5-1·7) | 1 | - | 1 |
| Heart failure | Hypertension | 392 | 3·6 (3·3-4·0) | 2·14 (1·91-2·40) | 2·06 (1·83-2·31) | 2·16 (1·89-2·47) |
|  | No hypertension | 1,098 | 1·8 (1·7-1·9) | 1 | 1 | 1 |
|  | QRISK2 ≥10% | 592 | 5·8 (5·3-6·3) | 4·02 (3·62-4·46) | - | 3·90 (3·49-4·34) |
|  | QRISK2 <10% | 898 | 1·5 (1·4-1·6) | 1 | - | 1 |
| Acute limb ischaemia | Hypertension | 30 | 0·3 (0·2-0·4) | 3·13 (2·01-4·89) | 3·00 (1·92-4·70) | 5·01 (2·99-8·38) |
|  | No hypertension | 56 | 0·1 (0·1-0·1) | 1 | 1 | 1 |
|  | QRISK2 ≥10% | 47 | 0·5 (0·3-0·6) | 7·34 (4·80-11·20) | - | 7·17 (4·65-11·07) |
|  | QRISK2 <10% | 39 | 0·1 (0·0-0·1) | 1 | - | 1 |
| Stroke or TIA | Hypertension | 450 | 4·1 (3·8-4·5) | 2·14 (1·92-2·38) | 2·09 (1·87-2·33) | 2·04 (1·80-2·30) |
|  | No hypertension | 1,250 | 2·1 (1·9-2·2) | 1 | 1 | 1 |
|  | QRISK2 ≥10% | 555 | 5·4 (5·0-5·9) | 2·95 (2·67-3·27) | - | 2·90 (2·61-3·22) |
|  | QRISK2 <10% | 1,145 | 1·9 (1·8-2·0) | 1 | - | 1 |
| CVD death | Hypertension | 173 | 1·6 (1·4-1·9) | 2·20 (1·85-2·62) | 2·12 (1·78-2·52) | 2·23 (1·81-2·75) |
|  | No hypertension | 472 | 0·8 (0·7-0·9) | 1 | 1 | 1 |
|  | QRISK2 ≥10% | 270 | 2·6 (2·3-3·0) | 4·39 (3·76-5·14) | - | 4·37 (3·68-5·18) |
|  | QRISK2 <10% | 375 | 0·6 (0·6-0·7) | 1 | - | 1 |

Total person-years per 1,000: hypertension = 108·6, no hypertension = 606·8, QRISK2 ≥10% = 102·4 and QRISK2 <10% = 613·0. LRT p-values all <0·001. *Hypertension models adjusted for: age, sex, ethnicity, socioeconomic status, BMI, alcohol intake and smoking status. QRISK2 models adjusted for: alcohol intake.

### **Table S9. Severe MACE after infection sensitivity analysis results**

| **Infection type** | **Cardiovascular risk** | **No. of events** | **Rate per 1,000 person-years (95% CI)** | **Crude**  **HR (95% CI)** | **Age and sex-adjusted**  **HR (95% CI)** | **Fully-adjusted***  **HR (95% CI)** |
| --- | --- | --- | --- | --- | --- | --- |
| ARI | Hypertension | 850 | 9·7 (9·0-10·4) | 2·11 (1·95-2·28) | 2·00 (1·85-2·16) | 2·02 (1·85-2·21) |
|  | No hypertension | 2,711 | 4·9 (4·7-5·1) | 1 | 1 | 1 |
|  | QRISK2 ≥10% | 1,321 | 15·2 (14·4-16·0) | 3·82 (3·57-4·09) | - | 3·71 (3·46-3·99) |
|  | QRISK2 <10% | 2,240 | 4·0 (3·9-4·2) | 1 | - | 1 |
| Influenza/ILI | Hypertension | 84 | 6·8 (5·5-8·5) | 2·50 (1·96-3·19) | 2·36 (1·85-3·03) | 2·37 (1·79-3·15) |
|  | No hypertension | 268 | 2·8 (2·5-3·2) | 1 | 1 | 1 |
|  | QRISK2 ≥10% | 96 | 9·6 (7·9-11·8) | 3·64 (2·88-4·60) | - | 3·59 (2·82-4·59) |
|  | QRISK2 <10% | 256 | 2·6 (2·3-3·0) | 1 | - | 1 |
| Pneumonia | Hypertension | 326 | 100·9 (90·2-113·3) | 1·65 (1·47-1·87) | 1·62 (1·43-1·83) | 1·65 (1·44-1·90) |
|  | No hypertension | 1,182 | 59·5 (56·1-63·1) | 1 | 1 | 1 |
|  | QRISK2 ≥10% | 544 | 124·1 (113·6-135·7) | 2·20 (1·98-2·44) | - | 2·15 (1·93-2·40) |
|  | QRISK2 <10% | 964 | 51·5 (48·3-55·0) | 1 | - | 1 |

*Hypertension models adjusted for: age, sex, ethnicity, socioeconomic status, BMI, alcohol intake and smoking status. QRISK2 models adjusted for: alcohol intake.

### **Table S10. Crude and adjusted incidence rate ratios for the association between cardiovascular risk and MACE after ARI among only patients who did not receive influenza or pneumococcal vaccine during follow-up**

| **Cardiovascular risk** | **No. of events** | **Rate per 1,000 person-years (95% CI)** | **Crude**  **HR (95% CI)** | **Age and sex-adjusted**  **HR (95% CI)** | **Fully-adjusted***  **HR (95% CI)** |
| --- | --- | --- | --- | --- | --- |
| Hypertension | 985 | 12·5 (11·8-13·3) | 2·17 (2·02-2·33) | 2·06 (1·92-2·22) | 2·07 (1·90-2·24) |
| No hypertension | 3,184 | 6·2 (5·9-6·4) | 1 | 1 | 1 |
| QRISK2 ≥10% | 1,526 | 20·2 (19·2-21·3) | 4·06 (3·81-4·32) | - | 3·96 (3·70-4·22) |
| QRISK2 <10% | 2,643 | 5·1 (4·9-5·3) | 1 | - | 1 |

Patients from 35,505 ARI episodes received influenza or pneumococcal vaccine. Among patients with raised cardiovascular risk, a higher proportion were vaccinated (hypertension=10% and QRISK2 ≥10%=13%) compared with low cardiovascular risk (no hypertension=6% and QRISK2 <10%=6%). None of the vaccinated patients had a MACE during follow-up. *Hypertension models adjusted for: age, sex, ethnicity, socioeconomic status, BMI, alcohol intake and smoking status. QRISK2 models adjusted for: alcohol intake.

### **References used in supplementary material**

1.	Levey AS, Stevens LA, Schmid CH, et al. A new equation to estimate glomerular filtration rate. *Ann Intern Med*. 2009;150(9):604-612. doi:10.7326/0003-4819-150-9-200905050-00006

2.	Hippisley-Cox J, Coupland C, Vinogradova Y, et al. Predicting cardiovascular risk in England and Wales: prospective derivation and validation of QRISK2. *BMJ*. 2008;336(7659):1475-1482. doi:10.1136/bmj.39609.449676.25

3.	National Health Service. Quality and Outcomes Framework (QOF). https://digital.nhs.uk/data-and-information/data-collections-and-data-sets/data-collections/quality-and-outcomes-framework-qof. Published 2020. Accessed March 3, 2020.
